# Genetics of Sleepwalking: Insights from whole exome sequencing

**DOI:** 10.1101/2025.09.17.25335508

**Authors:** Saiyet de la Caridad Baez Llovio, Yves Dauvilliers, Valerie Triassi, Véronique Daneault, Marjorie Labrecque, Simon Fournier, Lucie Barateau, Regis Lopez, Antonio Zadra, Simon C. Warby, Alex Desautels, Martine Tetreault

**Author notes:** These authors contributed equally to this work (Yves Dauvilliers, Alex Desautels, Martine Tetreault). Correspondence to: Martine Tetreault and Alex Desautels, Full address: - University of Montreal Hospital Research Centre (CRCHUM), Montréal, QC H2X 0A9, Canada. - Center for Advanced Research in Sleep Medicine, Sacré-Cœur Hospital, Montréal, QC, Canada. and.

## Abstract

Sleepwalking (SW) is a sleep disorder that belongs to the non-rapid eye movement (NREM) sleep family of parasomnias. Although linkage analyses in large families suggest that some forms of SW may follow a monogenic inheritance pattern, the genetic basis of SW has not been thoroughly investigated. The objective of this study was to investigate the role of rare genetic variants in sleepwalking by performing whole-exome sequencing (WES) in two independent cohorts.

WES was performed on a cohort of 254 individuals diagnosed with SW (54.7% female, mean age: 39.1 ± 10.7 years) and 124 control individuals were selected based on age and sex (52.4% female, all aged 18 years or older), from Montreal, Canada and Montpellier, France. To be included in the SW group, probands were required to have a primary complaint of SW, undergone at least one night of video-polysomnography, and to experience at least one parasomnia episode per month. By focusing on rare, potentially deleterious genetic variants, defined as having a minor allele frequency (MAF) ≤ 5% and a Combined Annotation Dependent Depletion (CADD) pathogenicity score ≥ 15, WES allowed us to detect novel contributors to the disorder that might be missed in studies focused on common variants.

We first identified 99 genes significantly enriched in patients with SW compared to the control group, with 92 genes overlapping between the two clinical cohorts. By prioritizing genes expressed in the brain, we found a strong genetic overlap between the two populations, with 31 genes carrying rare variants in common, including the top 10 genes with the highest contribution to SW compared to controls: *NPIPB13, SRRM2, SIRT1, CANT1, DPYSL5, ABCC10, ELF2, DPP9*, *RBM28* and *MCF2L2*. Results were validated using an independent control cohort from the CARTaGENE database, except for *MCF2L2*. The genes *NPIPB13*, *SRRM2*, and *SIRT1* displayed the highest contributions in the population, with values of 13.1%, 7.5%, and 5.5%, respectively. This study represents an important step toward understanding the genetic architecture of sleepwalking, particularly the role of rare coding variants, in sleepwalking and opens new avenues for future research into the disorder’s underlying biological mechanisms.

## Introduction

Sleepwalking (SW), also known as somnambulism, is a sleep disorder classified as a non-rapid eye movement (NREM) parasomnia (ICSD-3 TR).^1^ SW is currently defined as recurrent episodes of incomplete awakening from sleep, with inappropriate or absent responsiveness, limited or no associated cognition or dream imagery, and partial or complete amnesia of the episode.^2^ SW typically occurs during the first third of the night, when slow-wave sleep (SWS) is most prevalent. During these episodes, individuals exhibit minimal responsiveness to their surroundings and display an electroencephalogram (EEG) reflecting both sleep-like and wake-like patterns.^3,4^ The widespread belief that sleepwalking is a benign condition is inaccurate. While childhood somnambulism is often transitory and relatively harmless, sleepwalking in adulthood carries considerable risks including episodes resulting in serious injuries to the sleeper or bed partner.^5,6^ In fact, somnambulism constitutes one of the leading causes of reported sleep-related injuries and forensic outcomes.^7–9^

Despite more than five decades of clinical and laboratory investigations, the pathophysiology of SW remains poorly understood. Unlike rapid eye movement (REM) sleep parasomnia, there is no animal model of NREM parasomnia. Human studies have consistently shown a strong genetic predisposition to the disorder.^10–14^ The familial nature of SW is largely recognized in up to 60% of cases yet the underlying mode of inheritance has not been elucidated. The rate of childhood SW increases with the number of affected parents: 22% when neither parent is affected, 45% when one parent is affected, and 60% when both are affected.^11,14^ Twin studies further support a genetic contribution, showing that adult SW is 5.3 times more common among monozygotic twins compared to dizygotic twins.^10^ Exploratory genetic analyses have identified a risk locus on chromosome 20q12-q13.12 in a single large family, as well as on chromosome 18 in individuals of African American descent, although no specific genes have yet been identified.^13,15^ Two other studies suggested a high prevalence of the HLA DQB1*05 allele and to a lesser extent DQB1*04 allele in sleepwalking and other types of NREM parasomnias.^12,16^

To date, only a few molecular studies have explored the genetic underpinnings basis of SW. We hypothesize that genetic factors play a significant role in the development of SW, with rare monogenic forms contributing to its aetiology. We aim to explore their role through whole-exome sequencing (WES) focusing on rare and likely pathogenic variants in two populations of SW compared to controls from Montreal (Canada) and Montpellier (France), and to validate results in an independent population of control subjects from the CARTaGENE database.

## Materials and methods

### Participants

Participants (patients with SW and controls) were recruited at the Center for Advanced Research in Sleep Medicine in Montreal, Canada and at the Sleep Disorders Unit in Montpellier, France. All patients had a primary complaint of SW. Patients had a clinical interview with a sleep expert to confirm the clinical characteristics (parasomnia subtype including SW, sleep terror and confusional arousals), frequency of SW episodes, age at onset, first- and second-degree family history of SW, and the presence of comorbidities. Each patient with SW underwent at least one-night video-polysomnography study. Inclusion criteria were: 1) typical clinical history of SW according to the ICSD3-TR,^1^ and 2) at least one parasomniac episode per month. Exclusion criteria were: positive clinical history of neurological diseases (such as epilepsy and parkinsonism), concomitant psychiatric disorder (i.e., depression, anxiety disorder, or psychotic disorder), central hypersomnolence disorder, intake of medication known to influence sleep architecture, and apnea-hypopnea index (AHI) > 10/h or an index of periodic leg movements during sleep > 15.

The age of sleepwalking onset was categorized as follows: “Childhood” (< 12 years old), “Teen” (12 – 17 years old) and “Adulthood” (≥ 18 years old). The frequency of episodes was defined as “Frequent”: one to two events or more per week and “Infrequent”: less than one event per week. Severity of SW was defined as “Severe” (i.e. with injury), “Moderate”, and “Mild”, according to a clinician-rated assessment based on the spectrum of nocturnal behaviors, ranging from elementary motor activity confined to bed, to complex ambulatory behaviors. Family history of sleepwalking among first- and second-degree relatives was self-reported and collected. Video-polysomnography was performed to rule out differential diagnoses such as sleep-related hypermotor epilepsy, REM Sleep Behavior Disorder (RBD) or sleep-related dissociative disorders, parasomnia behaviors during slow-wave sleep and confirm the occurrence of SWS interruptions as previously described.^17^ The presence of comorbidities such as insomnia (any type), sleep apnea, or Restless Legs Syndrome (RLS) was recorded in patients.

Drug-free control participants were selected based on age, sex, and recruitment center (i.e., the Center for Advanced Research in Sleep Medicine in Montréal, Canada, and the Sleep Disorders Unit in Montpellier, France). Controls had no current or history of sleepwalking, other parasomnias, neurological disorders, or psychiatric conditions. Information on comorbidities including any form of insomnia (any type), sleep apnea, RLS, or autoimmune diseases was also collected. All participants provided a written informed consent prior to study enrollment, and the study protocols were approved by the respective institutional review boards.

### DNA extraction, preparation and quantification

DNA extraction and preparation were performed in standardized conditions for all participants. DNA was extracted from all participants at Sacré-Coeur Hospital from buffy coat samples using the FlexiGene DNA Kit 250 (Qiagen), following the manufacturer’s guidelines. DNA concentration and purity were determined with UV spectrophotometry.

### Bioinformatics

WES was conducted at Centre d’Expertise et de Services Genome Quebec using SureSelect Human Exome Library (Agilent) on a NovaSeq6000 (Illumina). Raw paired-end sequencing files (FASTQ) were processed using our in-house pipeline as previously described,^18^ with alignment to the hg19 reference genome performed using Burrows-Wheeler Aligner (BWA). Variant calling was done using VarDict tool, followed by annotation through the ANNOVAR pipeline. For variant filtering, we focused on rare variants with a minor allele frequency (MAF) ≤LJ5% in the gnomAD (Genome Aggregation Database) and Combined Annotation Dependent Depletion (CADD) score ≥LJ15, indicating potential functional impact.^19^ Given that many variants were unique to individual samples, we employed a gene-based approach for downstream analysis. A Fisher’s exact test was conducted to compare the distribution of variant counts between the entire SW population and the control group, as well as within each country independently. The analysis was conducted using NumPy, SciPy, and Statsmodels Python packages. Enrichment analysis was performed in R to compare gene frequency between SW cases and controls for each gene, using the grepel and ggplot2 packages as well as custom scripts. Variants identified as heterozygous were assigned a value of 1, and those identified as homozygous were assigned a value of 2, representing the number of occurrences.

To assess gene expression in the brain, we primarily utilized the GTEx database (https://www.gtexportal.org/home/), supplemented by data from the Protein Atlas (https://www.proteinatlas.org) and GeneCards (https://www.genecards.org). Gene expression levels were measured in Transcripts Per Million (TPM), an arbitrary unit used for normalization providing a relative measure of transcript abundance. Genes were considered “brain-expressed” if they exhibited TPM > 25 across multiple brain regions.^20^

To assess the contribution score, we first identified the chromosomal location of each variant and quantified its occurrence within each cohort. Variants that were enriched in the control cohort were excluded from further analysis. Contribution scores were calculated as the proportion of variant occurrences in the sleepwalking cohort relative to the total number of alleles, expressed as a percentage. Variants identified in the same gene across multiple individuals were counted only once.

### CARTaGENE Database (CaG)

Our results were compared in an independent population of control subjects from the CARTaGENE database, a public database in Quebec that contains data on the health and lifestyle of Quebec men and women between the ages of 40 and 69. From this cohort participants were selected based on the availability of whole-exome sequencing data, their health status, and the absence of sleep-related complaints. This selection resulted in a control group of 198 individuals.^21^

## Results

### Population characteristics

Based on the exclusion criteria, 254 patients with primary SW (129 from Montreal, Canada, and 125 from Montpellier, France) were included in the analysis. Among them, 139 were female patients representing 54.7% of the total, with an average age of 39.1 ± 10.7 years. Twenty-six subjects were younger than 18 years of age at the time of the study. The male-to-female ratios were 1:1.5 (52:77) and 1:1 (63:62), in the population from Montréal and from Montpellier respectively (see Table 1). A family history of SW was reported in 49.6% of participants (47.3% in Montreal and 52% in Montpellier). For 71.2% of participants, SW episodes began in childhood, 17% reported the onset during adolescence, and 11.8% in adulthood. Overall, 92.9% of patients were classified as having moderate SW severity, while 84.6% had frequent episodes. Comorbidities were present in 16.5% of cases with 14% having insomnia, 11.6% sleep apnea, and 7.2% RLS. Detailed information about the patients can be found in Supplementary Table 1.

**Table 1:** Demographic and clinical characteristics of Study cohort.

| Cohort | Sleepwalking |  | Controls |  |
| --- | --- | --- | --- | --- |
|  | Montréal | Montpellier | Montréal | Montpellier |
| N (378) | 129 | 125 | 64 | 60 |
| Age (Mean ± SD) | 34 ± 11.40 | 27 ± 10.70 | ≥ 18 | ≥ 18 |
| Sex (Male:Female ratio) | 52:77 | 63:62 | 28:36 | 31:29 |
| Family History (Yes:No ratio) | 47.30% - 45.70% (7%NA) | 52% - 44% (4%NA) | - | - |
| Age of onset (Child/Teen/Adult) | 65.10% - 18.60% - 16.30% | 77.60% - 15.20% - 7.20% | - | - |
| Severity (Mild/Moderate/Severe) | 0.80% - 87.60% - 11.60% | 0 - 98.40% - 1.60% | - | - |
| Frequency of the Episodes (Frequent/Infrequent) | 76% - 18% (6%NA) | 93.60% - 0.80% (5.60%NA) | - | - |
| Comorbidities (Yes/No) | 25.50% - 74.50% | 7.20% - 92.80% | 98.40% - 1.60% | 38.40% - 61.60% |
| Comorbidities classification: Insomnia/Sleep apnea/Restless Legs Syndrome (RLS)/Autoimmune diseases | 14% - 11.60% - 0 - 0 | 0 - 0 - 7.20% - 0 | 73.50% - 9.30% - 17.20% - 0 | 25% - 0 - 1.60% - 11.60% |

The control group consisted of 124 individuals aged 18 years or older, with 64 from Montréal and 60 from Montpellier. Women comprised 52.4% of the group. Comorbidities were identified in 60.5% of the control group, including 49.2% with insomnia, 4.8% with sleep apnea, 9.8% with RLS, and 5.7% with autoimmune diseases. Detailed information about the control group can be found in Supplementary Table 2.

### WES analysis reveals genes significantly enriched in the SW individuals

To evaluate the contribution of rare variants to SW, enrichment analysis was performed comparing patients and controls, focusing on rare, likely pathogenic variants. We identified 99 genes significantly enriched (p < 0.05) in patients with SW compared to the control group (Figure 1A and Supplementary Table 3). Most of these genes carried multiple variants, resulting in a total of 204 variants. Interestingly, 92 genes were commonly shared between the Montreal and Montpellier SW cohorts, indicating a strong overlap in genetic enrichment.

**Figure 1:**
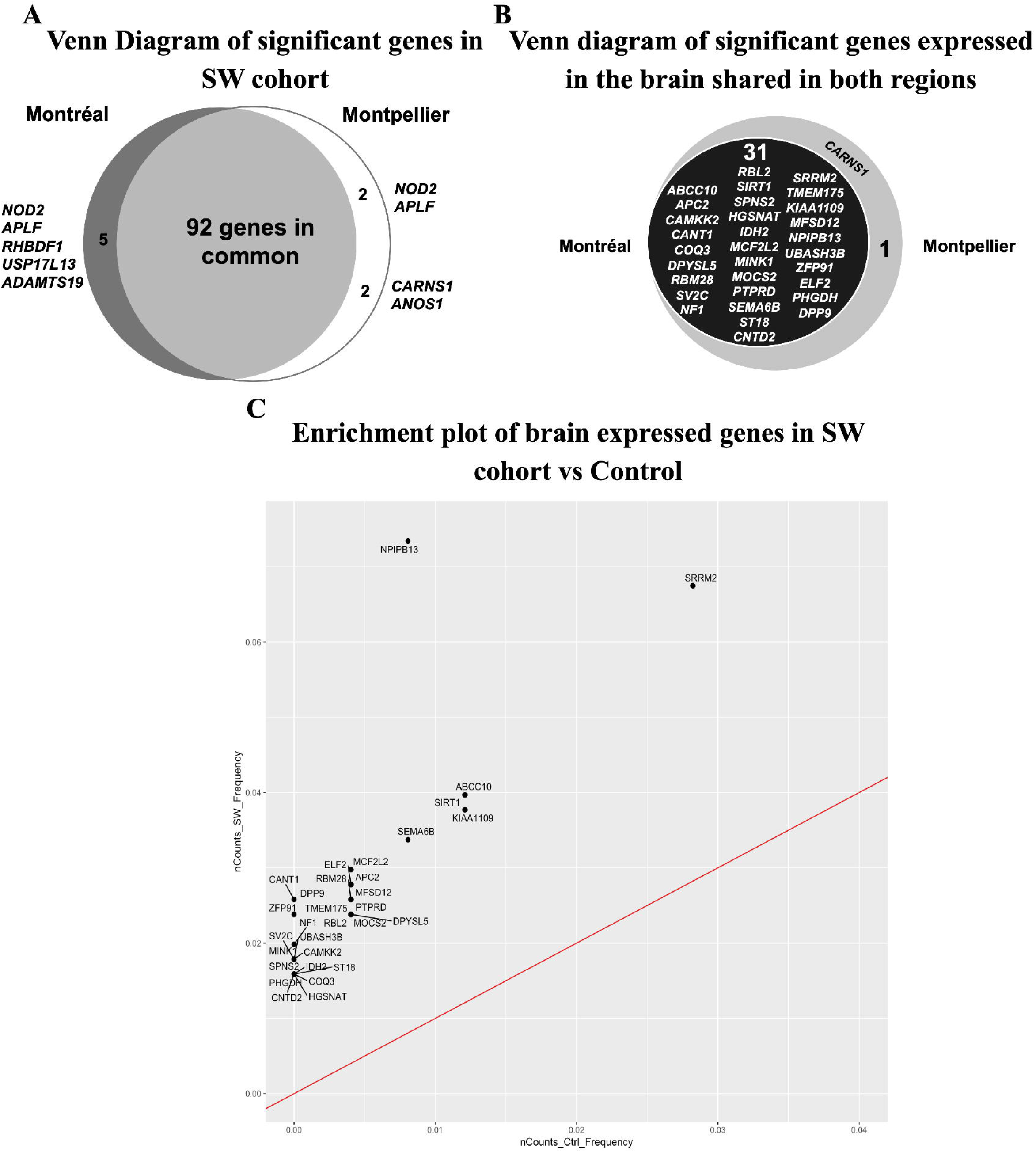
Genes significantly enriched in the SW cohort. **(A)** Venn diagram showing Fisher test significant genes p-value < 0.05. (**B)** Venn diagram showing 32 Fisher test significant genes expressed in the brain shared between Montreal and Montpellier regions (Transcripts Per Million, TPM >25). (**C)** Enrichment plot of the significant brain expressed genes in SW cohort compared to controls. The X-axis represents the allelic frequencies in healthy controls, while the Y-axis shows the allelic frequencies in SW patients, both expressed as relative frequencies. 31 significant genes were found to have a higher allelic frequency in SW patients compared to the control group. The red line represents the point of equal allelic frequencies between the two groups, serving as a reference for comparison.

When compared to their respective control group in each population, *RHBDF1*, *USP17L13*, and *ADAMTS19* were uniquely associated with the SW cohort in Montreal, while *CARNS1* and *ANOS1* were specific to the SW cohort in Montpellier. Although *APLF* and *NOD2* variants were enriched in the population of SW from Montreal, they were absent in the Montpellier SW cohort but reported with a high frequency in the Montpellier control group, suggesting they are unlikely to be involved in the aetiology of sleepwalking.

In relation to the pathophysiological nature of sleepwalking, using our significantly enriched genes (99 genes) we refined our analysis by prioritizing genes based on their expression in the brain. This approach resulted in a reference list consisting of 31 genes (75 variants) common to both the Montreal and Montpellier cohorts (Figure 1B). An enrichment plot of brain-expressed genes revealed a higher frequency of these genes in the SW cohort compared to the control cases (Figure 1C), which is indicative of their potential relevance to SW condition. We then evaluated the contribution of these 31 genes within the SW cohort and identified the top 10 genes based on their contribution percentages (Table 2 and Supplementary Table 4). The genes *NPIPB13*, *SRRM2*, and *SIRT1* displayed the highest contribution percentages in the population, with values of 13.1%, 7.5%, and 5.5%, respectively. All variants were heterozygous, except for one homozygous variant (rs1240720619), which showed low sequencing coverage. When analyzing the contribution of these genes to potential familial cases, we found that 27% of probands from familial forms carried variants in *NPIPB13*, 15.6% in *SRRM2*, and 11.4% in *SIRT1*.

**Table 2:** Table of the top 10 genes based on their contributions to the SW cohort.

| Gene | Chr | Position | rsID | Number of counts in controls | Number of counts in SW | Contribution score (% in population) | Contribution to familial cases (%) |
| --- | --- | --- | --- | --- | --- | --- | --- |
| NPIPB13 | 16 | 30236217 | rs796810494 | 0 | 32 | 13.10 | 27 |
|  |  | 30237003 | rs1240720619 | 0 | 1 |  |  |
| SRRM2 | 16 | 2814476 | rs1438787177 | 0 | 2 | 7.56 | 15.60 |
|  |  | 2814911 | rs149555495 | 0 | 3 |  |  |
|  |  | 2816519 | rs138447860 | 0 | 6 |  |  |
|  |  | 2816627 | rs117133016 | 1 | 8 |  |  |
|  |  | 2817667 | rs149458303 | 0 | 2 |  |  |
| SIRT1 | 10 | 69644532 | - | 0 | 10 | 5.54 | 11.40 |
|  |  | 69672479 | rs116040871 | 0 | 4 |  |  |
| CANT1 | 17 | 76989666 | rs34082669 | 0 | 4 | 4.78 | 9.87 |
|  |  | 76989715 | rs139486406 | 0 | 3 |  |  |
|  |  | 76991115 | rs772754957 | 0 | 3 |  |  |
|  |  | 76993649 | rs144060377 | 0 | 2 |  |  |
| DPYSL5 | 2 | 27121449 | rs139435744 | 0 | 2 | 4.38 | 9 |
|  |  | 27157542 | rs151073506 | 1 | 9 |  |  |
| ABCC10 | 6 | 43400205 | rs144459262 | 1 | 8 | 4.38 | 9 |
|  |  | 43400850 | rs150124239 | 0 | 3 |  |  |
|  |  | 43406366 | rs142854084 | 0 | 3 |  |  |
| MCF2L2 | 3 | 182923984 | rs61750384 | 0 | 2 | 4 | 8.26 |
|  |  | 182946129 | rs61753470 | 0 | 3 |  |  |
|  |  | 183006997 | rs201525425 | 0 | 2 |  |  |
|  |  | 183017837 | rs144274760 | 0 | 3 |  |  |
| ELF2 | 4 | 139981622 | rs147539143 | 0 | 2 | 3.98 | 8.22 |
|  |  | 139993175 | rs17322140 | 0 | 8 |  |  |
| DPP9 | 19 | 4685696 | rs762729074 | 0 | 2 | 3.96 | 8.18 |
|  |  | 4689635 | rs200229921 | 0 | 4 |  |  |
|  |  | 4704192 | rs200878232 | 0 | 4 |  |  |
| RBM28 | 7 | 127953238 | rs73230638 | 1 | 6 | 3.96 | 8.18 |
|  |  | 127963597 | rs138703329 | 0 | 4 |  |  |

In the *NPIPB13* gene, two variants were identified: rs796810494 and rs1240720619. The rs796810494 variant was found in 32 individuals with SW, including 27 from Montreal and 5 from Montpellier, and none were observed in the control groups (Table 2). In contrast, the rs1240720619 variant was detected exclusively in one individual from the Montreal cohort. When stratifying the phenotype by these variants, we observed that 57.5% of carriers had a familial history of SW, and that 60.6% of them experienced the onset of symptoms during childhood. Additionally, 87.8% of them reported frequent episodes, and exhibited moderate SW severity.

For the *SRRM2* gene, five variants were identified: rs117133016, rs138447860, rs14955549, rs1438787177, and rs149458303. The rs117133016 variant was observed in 8 cases (6 from Montreal and 2 from Montpellier) and in only one control case, while rs138447860 was present in 6 cases (4 from Montreal and 2 from Montpellier) but not in controls. The rs14955549 variant was detected exclusively in 3 individuals from Montreal, whereas rs1438787177 and rs149458303 were each identified in one case from Montreal and one from Montpellier. Overall, 57.8% of individuals had a familial history of SW, and 73.7% developed the condition in childhood. Additionally, 89.5% reported frequent episodes, and 78.9% exhibited moderate severity.

Two variants were identified in the *SIRT1* gene: chr10:69644532 and rs116040871. The chr10:69644532 variant was present in 10 cases, 6 individuals from Montreal and 4 from Montpellier, while rs116040871 was detected in 4 cases (3 from Montpellier and 1 from Montreal), and none in the controls. Among these individuals, 42.8% had a family history of sleepwalking, and 64.2% experienced early-onset symptoms. Notably, all cases reported frequent episodes, and 85.7% exhibited moderate severity.

The remaining genes in the top 10 list, based on their contribution percentages, include *CANT1, DPYSL5, ABCC10, MCF2L2, ELF2, DPP9*, and *RMB28* (Table 2). To investigate whether SW patients carried multiple rare variants across the top 10 genes previously identified, we manually examined each gene to determine which samples harbored rare variants. A total of 121 subjects carried rare variants in the top 10 genes ranked by contribution (Table 2). Among them, 24 individuals (19.8%) harbored two or more variants. Specifically, 15 individuals (12.4%) carried two variants, 8 (6.7%) carried three, and 1 individual (0.8%) carried four variants. We examined the co-occurrence of specific variants and did not detect any consistent co-occurrence patterns. However, at the gene level, the most frequent co-occurrence was between *SIRT1* and *SRRM2*, observed in four individuals. Supplementary Table 5 summarizes the number of rare variants per sample across these genes. We also investigated whether the combination of rare variants in different genes might contribute to disease risk. Such effects could act additively on biological pathways or processes relevant to the phenotype.^23^ However, no consistent pattern emerged, suggesting that the co-occurrence of variants does not support a synergistic pathogenic effect.

### Validation of WES results in an independent control group: CARTaGENE (CaG) cohort

To validate our findings, we compared the panel of 31 SW enriched genes from our previous analysis against an independent control group from the CaG database. An enrichment plot revealed a higher prevalence of 28 brain-expressed genes in SW patients compared to controls, along with higher allelic frequency in the SW cohort, as illustrated in Figure 2. Notably, *NPIPB13*, *SRRM2*, *SIRT1*, and *ABCC10* exhibited the highest allele frequency values from our top10 list and thus demonstrates a concordance between our WES results and those of an entirely different control group. In contrast, only *APC2*, *COQ3*, and *MCF2L2* were not enriched in SW relative to CaG controls, with *MCF2L2* being the only one from the top10 list. This finding suggests that these genes are unlikely to play a major role in the development of SW.

**Figure 2:**
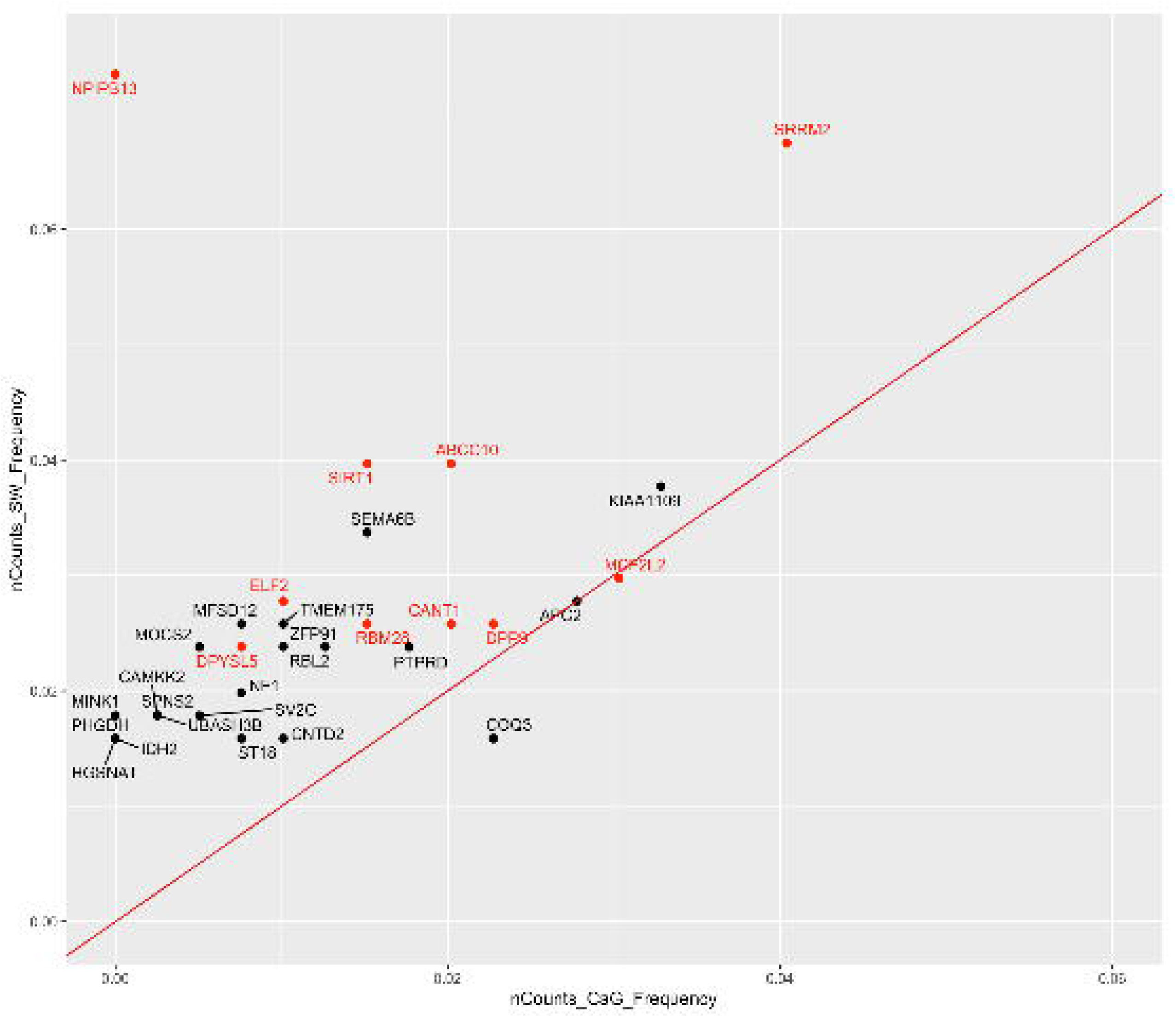
Genes significantly enriched in the SW cohort compared to CARTaGENE (CaG) cohort. Enrichment plot of the significant brain expressed genes in SW cohort compared to CARTaGENE (CaG) control database. The X-axis represents the allelic frequencies in healthy controls, while the Y-axis shows the allelic frequencies in SW patients, both expressed as relative frequencies. 28 significant genes were found to have a higher allelic frequency in SW patients compared to the control group, while 3 of them *APC2*, *COQ3* and *MCF2L2* have a higher allelic frequency in controls. The red line represents the point of equal allelic frequencies between the two groups, serving as a reference for comparison.

## Discussion

Based on multiple lines of evidence supporting a genetic predisposition to sleepwalking, this study explored potential connections at the gene level using WES. We observed a strong genetic overlap between two independent sleepwalking cohorts from Montreal, Canada and Montpellier, France, suggesting a shared genetic background across these populations. Our findings indicate that the 92 significant genes identified may contribute to SW susceptibility and may also reflect population-specific genetic traits, further supporting the idea of a shared genetic cause for the condition. A deeper examination of the significant brain-expressed genes in the SW cohort reveals important insights into their genetic contributions, highlighting the top 10 genes with potential involvement in SW. Using CaG as an independent validation cohort strengthened the reliability and generalizability of our findings, with the overall results supporting the involvement of 9 candidate genes in SW.

Previous studies have identified genetic loci associated with SW. They investigated the inheritance patterns and reported a risk locus on chromosome 20q12–13.12,^13^ while another study conducted a genome-wide association study (GWAS) and found an association on chromosome 18 in individuals of African American descent, but not in those of European descent.^15^ Our study did not replicate these findings, most probably due to a focus on the rare variants, lack of genetic data on family members and differences in sample size. The differences between previous studies and our findings might also be attributed to differences in population, genetic ancestry, and inclusion criteria. However, we identified chromosome 16 as a “hot-spot” for rare potential SW-related genes, containing 10 significant genes (*NPIPB13*, *SRRM2*, *RBL2*, *RPL3L*, *ZSCAN10*, *RHBDF1*, *BICDL2*, *ERCC4*, *GLIS2*, *NOD2*). Additionally, our study did not identify any rare variants in the HLA genes, based on the established filtering criteria, that could be linked to SW. It’s important to note that the HLA genes are highly polymorphic, and their variants are typically classified as common rather than rare,^24^ which may explain this result.

The high prevalence of familial history in our cohort supports the hypothesis of a genetic basis for SW. In our study, *NPIPB13*, *SRRM2*, and *SIRT1* have emerged as the most promising candidate genes. More than half of the genetic contribution in familial cases can be attributed to variants in these 3 genes: the *NPIPB13* gene representing 27% of familial cases, followed by *SRRM2* and *SIRT1* contributing 15.6% and 11.4%, respectively. However, familial cases were self-reported by the probands, and the absence of DNA samples from potentially affected relatives limits our ability to perform segregation analyses, which are crucial for confirming the inheritance patterns and pathogenicity.

For further discussion, we focused on the nine genes that were consistently validated in both cohorts.

*NPIPB13* is a member of the nuclear pore complex interacting protein family with a central role in structuring this complex, facilitating the translocation of molecules.^25^ Interestingly, *NPIPB13* is located within the 16p11.2 locus. The human 16p11.2 microdeletion is one of the most common chromosome copy number variations (CNVs) associated with neurodevelopmental disorders;^26^ this region has also been associated with sleep disturbances in humans ^27,28^ and animal models.^29^ There is no animal model of SW; however, mouse models of human 16p11.2 microdeletion have demonstrated pathophysiological impairments in synaptic transmission and local circuits in the hippocampus^29^ and striatum.^30^ Other groups demonstrated that these mice have altered sleep architecture, with increased wake time, reduced REM and NREM sleep, and deficient NREM–REM transitions.^29^ One of the most prominent characteristics of sleepwalking is the disruption of NREM sleep continuity, as evidenced by an increased frequency of spontaneous awakenings and EEG arousals during SWS.^3,4,17,31^ Additionally, *NPIPB13* is highly expressed in the cerebellum.^32^ While more research is necessary to establish the cerebellum’s role in SW, some studies have indicated increased activity in the anterior cerebellum during these episodes.^33^ All these findings suggest that *NPIPB13* may play a role in the motor behaviors associated with SW.

*SRRM2* (OMIM ID: *606032) is a key regulator of mRNA splicing. Although it has not been previously associated with sleep conditions, variants in *SRRM2* have been linked to neurodevelopmental disorders.^34,35^ Clinical features observed in individuals with *SRRM2* variants include developmental delay, attention deficit hyperactivity disorder (ADHD), hypotonia, gastroesophageal reflux, obesity, and autism.^35^

*SIRT1* (OMIM ID: *604479) encodes a NAD+-dependent deacetylase that plays a fundamental role in regulating the circadian clock and maintaining telomere homeostasis.^36,37^ *SIRT1* is essential for sleep-wake regulation, and its reduced activity or gene deletion have shown to disrupt the circadian cycle.^38^ It regulates the acetylation of clock components and the circadian amplitude of clock genes by mediating *CLOCK/BMAL1* complex formation, promoting *BMAL1* transcription, and facilitating *PER2* degradation; however its involvement in sleep architecture remains unknown.^36^

*ABCC10* is a member of the ABC transporter family. These transporters are widely known for their involvement in regulating molecular transport by expelling substances back into the bloodstream, restricting their entry into the central nervous system.^39^ Their expression in healthy tissues follows a circadian rhythm, with its activity varying depending on the time of day.^40^

*CANT1*, *DPYSL5*, and *RBM28* have been previously associated with neurodevelopmental disorders, including developmental delays, progressive neurological impairments, and severe intellectual disabilities.^41–43^ Studies have shown that individuals with such conditions, especially children, frequently experience sleep disturbances^44,45^ making this an important factor to consider in the context of SW.

*DPP9* is a member of the dipeptidyl peptidase IV family. Although its exact biological function remains unclear, recent studies have shown that inhibition of *DPP9* can activate the NLRP1 inflammasome, suggesting a role in immune regulation during infections.^46^ In addition, *DPP9* has been implicated in memory formation through modulation of the endopeptidase pathway and enhancement of synaptic plasticity.^47^

Finally, *ELF2*, also known as *NERF*, is a transcription factor belonging to the Erythroblast Transformation Specific (ETS) family. It plays a key role in regulating genes involved in B and T cell development, cell cycle progression, and angiogenesis.^48^ Recent findings have identified *ELF2* gene variants in individuals with Cerebellar Ataxia, Neuropathy, and Vestibular Areflexia Syndrome (CANVAS),^49^ further supporting its neurological relevance and suggesting a potential link with the cerebellum.

We also examined here whether the combination of rare variants from the different genes contributes to SW for these patients but failed to detect a consistent pattern. However, gene-gene interactions cannot be ruled out, as the effect of one variant may depend on the presence of another in a different gene and moreover little is known about certain genes. A potential area for further investigation is to determine if the observed variants act independently or influence each other’s effects, specifically for patients carrying multiple rare variants. They can contribute to different phenotypes and related clinical features, such as earlier onset, or a stronger family history, which may indicate the presence of a distinct subgroup within the SW population.

Our study provides a prioritized list of candidate genes potentially involved in SW. However, since functional validation in animal models is currently not feasible, replication in larger and independent cohorts is essential to confirm these findings and strengthen the evidence for their involvement. Despite our small sample size in comparison to genetic studies of other disorders, this remains the largest cohort studied to date for the genetics of SW. Genetics has been implicated as a contributing factor in SW, and the use of three independent control cohorts (Montreal, Montpellier, and CaG) of comparable ethnic origin adds strength to our study. Identifying the genes contributing to SW is crucial for advancing our understanding of its aetiology, diagnosis, and treatment.

In conclusion, our findings reveal significant rare genetic variants shared between two distinct populations: Montreal, Canada and Montpellier, France, highlighting a notable genetic overlap and suggesting the presence of rare genetic factors contributing to SW. This points to the existence of susceptibility genes that may play a causal role in the development of this pathology. Future research should aim to uncover additional common variants that could explain unresolved cases, validate these findings in larger and more diverse independent cohorts, and further elucidate the genetic mechanisms underlying SW.

## Data availability

Exome data have been deposited in the Gene Expression Omnibus (GEO) database as GSExxxxxx

## Acknowledgments

The authors would like to acknowledge the patients who participated in this study. We also thank the Centre d’Expertise et de Services Génome Québec for their support, and the editors for reviewing the manuscript. We are grateful to the Digital Research Alliance of Canada for providing computing resources (Beluga), and to CARTaGENE for access to data.

## Abbreviations

ADHD: Attention Deficit Hyperactivity Disorder
AHI: Apnea Hypopnea Index
BWA: Burrows-Wheeler Aligner
CADD: Combined Annotation Dependent Depletion
CaG: CARTaGENE
CANVAS: Cerebellar Ataxia, Neuropathy, and Vestibular Areflexia Syndrome
CNV: Copy Number Variants
DNA: Deoxyribonucleic acid
EEG: Electroencephalogram
ETS: Erythroblast Transformation Specific
GEO: Gene Expression Omnibus
gnomAD: Genome Aggregation Database
GWAS: Genome-Wide Association Study
MAF: Minor Allele Frequency
NREM: Non-Rapid Eye Movement
RBD: REM Sleep Behavior Disorder
REM: Rapid Eye Movement
RLS: Restless Legs Syndrome
SW: Sleepwalking
SWS: Slow-Wave Sleep
TPM: Transcripts Per Million
WES: Whole-Exome Sequencin

## Supplementary material

Supplementary material is available at Brain online

## Supplementary material

**Supplementary Table 1: Detailed clinical information of each individual in the SW cohort.**

**Supplementary Table 2: Detailed clinical information of each individual in the Control cohort.**

**Supplementary Table 3: Fisher’s Exact Test Results for SW Case-Control Comparison.**

The table shows the 99 genes significantly enriched (p < 0.05) with the Odds Ratios and 95% Confidence Intervals.

**Supplementary Table 4: Contribution of Enriched Genes in the SW cohort.**

The table shows the number of counts of the significantly enriched genes in the SW cohort compared to the Control cohort. For each gene/variant the table includes the corresponding contribution scores and brain expression level.

**Supplementary Table 5: Table of individuals in the SW cohort carrying two or more variants within the top 10 genes.**

The table shows the number of rare variants per sample across these genes

## References

1. American Academy of Sleep Medicine. International Classification of Sleep Disorders, 3rd Edn. 3rd ed. American Academy of Sleep Medicine; 2014.

2. Arnulf I. Sleepwalking. Current Biology. 2018;28(22):R1288-R1289. doi:10.1016/j.cub.2018.09.062

3. Castelnovo A, Lopez R, Proserpio P, Nobili L, Dauvilliers Y. NREM sleep parasomnias as disorders of sleep-state dissociation. Nat Rev Neurol. 2018;14(8):470–481. doi:10.1038/s41582-018-0030-y

4. Lopez R, Dauvilliers Y. Challenges in diagnosing NREM parasomnias: Implications for future diagnostic classifications. Sleep Med Rev. 2024;73:101888. doi:10.1016/j.smrv.2023.101888

5. Moldofsky H, Gilbert R, Lue FA, Maclean AW. Sleep-Related Violence. Vol 18.; 1995. https://academic.oup.com/sleep/article/18/9/731/2749722

6. Schenck CH, Milner DM, T D Hurwitz TD, Bundlie SR, Mahowald MW. A polysomnographic and clinical report on sleep-related injury in 100 adult patients. American Journal of Psychiatry. 1989;146(9):1166–1173. doi:10.1176/ajp.146.9.1166

7. Correa VM, Arruda GC, Szűcs A. Parasomnia patients and risk of injury, a 16-years clinical study. Sleep Epidemiology. 2023;3:100057. doi:10.1016/j.sleepe.2023.100057

8. Lopez R, Jaussent I, Scholz S, Bayard S, Montplaisir J, Dauvilliers Y. Functional Impairment in Adult Sleepwalkers: A Case-Control Study. Sleep. 2013;36(3):345–351. doi:10.5665/sleep.2446

9. Pressman MR. Factors that predispose, prime and precipitate NREM parasomnias in adults: Clinical and forensic implications. Sleep Med Rev. 2007;11(1):5–30. doi:10.1016/j.smrv.2006.06.003

10. Hublin C, Kaprio; J, Partinen; M, Heikkila; K, Koskenvuo M. Prevalence and Genetics of Sleepwalking: A Population-Based Twin Study. Vol 48.; 1997.

11. Kales A, Soldatos CR, Bixler EO, et al. Hereditary Factors in Sleepwalking and Night Terrors. British Journal of Psychiatry. 1980;137(2):111–118. doi:10.1192/bjp.137.2.111

12. Lecendreux M, Bassetti C, Dauvilliers Y, Mayer G, Neidhart E, Tafti M. HLA and genetic susceptibility to sleepwalking. Mol Psychiatry. 2003;8(1):114–117. doi:10.1038/sj.mp.4001203

13. Licis AK, Desruisseau DM, Yamada BKA, Duntley SP, Gurnett CA. Novel Genetic Findings in an Extended Family Pedigree with Sleepwalking.; 2010. https://www.neurology.org

14. Petit D, Pennestri MH, Paquet J, et al. Childhood Sleepwalking and Sleep Terrors. JAMA Pediatr. 2015;169(7):653. doi:10.1001/jamapediatrics.2015.127

15. Chiba Y, Phillips OR, Takenoshita S, et al. Genetic and demographic predisposing factors associated with pediatric sleepwalking in the Philadelphia Neurodevelopmental Cohort. J Neurol Sci. 2021;430:119997. doi:10.1016/j.jns.2021.119997

16. Heidbreder A, Frauscher B, Mitterling T, et al. Not Only Sleepwalking But NREM Parasomnia Irrespective of the Type Is Associated with HLA DQB1*05:01. Journal of Clinical Sleep Medicine. 2016;12(04):565–570. doi:10.5664/jcsm.5692

17. Lopez R, Shen Y, Chenini S, et al. Diagnostic criteria for disorders of arousal: A video□polysomnographic assessment. Ann Neurol. 2018;83(2):341–351. doi:10.1002/ana.25153

18. Touma L, Labrecque M, Tetreault M, Duquette A. Identification and Classification of Rare Variants in NPC1 and NPC2 in Quebec. Sci Rep. 2021;11(1):10344. doi:10.1038/s41598-021-89630-5

19. Laflamme N, Triassi V, Martineau L, et al. X□Linked Bilateral Polymicrogyria With Epilepsy and Intellectual Disability Associated With a Novel <scp> *KIF4A* </scp> Variant. Am J Med Genet A. 2024;197(1). doi:10.1002/ajmg.a.63860

20. Vojinovic D, Kavousi M, Ghanbari M, et al. Whole-Genome Linkage Scan Combined With Exome Sequencing Identifies Novel Candidate Genes for Carotid Intima-Media Thickness. Front Genet. 2018;9. doi:10.3389/fgene.2018.00420

21. Awadalla P, Boileau C, Payette Y, et al. Cohort profile of the CARTaGENE study: Quebec’s population-based biobank for public health and personalized genomics. Int J Epidemiol. 2013;42(5):1285–1299. doi:10.1093/ije/dys160

22. Tétreault M, Duquette A, Thiffault I, et al. A new form of congenital muscular dystrophy with joint hyperlaxity maps to 3p23-21. Brain. 2006;129(Pt 8):2077–2084. doi:10.1093/brain/awl146

23. Li J, Kong N, Han B, Sul JH. Rare variants regulate expression of nearby individual genes in multiple tissues. PLoS Genet. 2021;17(6):e1009596. doi:10.1371/journal.pgen.1009596

24. Creary LE, Sacchi N, Mazzocco M, et al. High-resolution HLA allele and haplotype frequencies in several unrelated populations determined by next generation sequencing: 17th International HLA and Immunogenetics Workshop joint report. Hum Immunol. 2021;82(7):505–522. doi:10.1016/j.humimm.2021.04.007

25. Stelzer G, Rosen N, Plaschkes I, et al. The GeneCards Suite: From Gene Data Mining to Disease Genome Sequence Analyses. Curr Protoc Bioinformatics. 2016;54(1). doi:10.1002/cpbi.5

26. Kumar RA, KaraMohamed S, Sudi J, et al. Recurrent 16p11.2 microdeletions in autism. Hum Mol Genet. 2007;17(4):628–638. doi:10.1093/hmg/ddm376

27. Kamara D, Beauchaine TP. A Review of Sleep Disturbances among Infants and Children with Neurodevelopmental Disorders. Rev J Autism Dev Disord. 2020;7(3):278–294. doi:10.1007/s40489-019-00193-8

28. Vos N, Kleinendorst L, van der Laan L, et al. Evaluation of 100 Dutch cases with 16p11.2 deletion and duplication syndromes; from clinical manifestations towards personalized treatment options. European Journal of Human Genetics. 2024;32(11):1387–1401. doi:10.1038/s41431-024-01601-2

29. Lu HC, Pollack H, Lefante JJ, Mills AA, Tian D. Altered sleep architecture, rapid eye movement sleep, and neural oscillation in a mouse model of human chromosome 16p11.2 microdeletion. Sleep. 2019;42(3). doi:10.1093/sleep/zsy253

30. Portmann T, Yang M, Mao R, et al. Behavioral Abnormalities and Circuit Defects in the Basal Ganglia of a Mouse Model of 16p11.2 Deletion Syndrome. Cell Rep. 2014;7(4):1077–1092. doi:10.1016/j.celrep.2014.03.036

31. Zadra A, Desautels A, Petit D, Montplaisir J. Somnambulism: clinical aspects and pathophysiological hypotheses. Lancet Neurol. 2013;12(3):285–294. doi:10.1016/S1474-4422(12)70322-8

32. Sjöstedt E, Zhong W, Fagerberg L, et al. An atlas of the protein-coding genes in the human, pig, and mouse brain. Science (1979). 2020;367(6482). doi:10.1126/science.aay5947

33. Bassetti C, Vella S, Donati F, Wielepp P, Weder B. SPECT during sleepwalking. The Lancet. 2000;356(9228):484-485. doi:10.1016/S0140-6736(00)02561-7

34. Cuinat S, Nizon M, Isidor B, et al. Loss-of-function variants in SRRM2 cause a neurodevelopmental disorder. Genetics in Medicine. 2022;24(8):1774–1780. doi:10.1016/j.gim.2022.04.011

35. Regan□Fendt KE, Rippert AL, Medne L, et al. Retrospective identification of patients with *SRRM2* □related neurodevelopmental disorder in a single tertiary children’s hospital. Am J Med Genet A. 2023;191(8):2149–2155. doi:10.1002/ajmg.a.63302

36. Osum M, Serakinci N. Impact of circadian disruption on health; SIRT1 and Telomeres. DNA Repair (Amst). 2020;96:102993. doi:10.1016/j.dnarep.2020.102993

37. Ribeiro RFN, Pereira D, de Almeida LP, Silva MMC, Cavadas C. SIRT1 activation and its circadian clock control: a promising approach against (frailty in) neurodegenerative disorders. Aging Clin Exp Res. 2022;34(12):2963–2976. doi:10.1007/s40520-022-02257-y

38. Huang T, Zhang X, Qi L, et al. Daytime Dysfunction: Symptoms Associated with Nervous System Disorders Mediated by SIRT1. Biomedicines. 2024;12(9):2070. doi:10.3390/biomedicines12092070

39. Furtado A, Mineiro R, Duarte AC, Gonçalves I, Santos CR, Quintela T. The Daily Expression of ABCC4 at the BCSFB Affects the Transport of Its Substrate Methotrexate. Int J Mol Sci. 2022;23(5):2443. doi:10.3390/ijms23052443

40. Kara Z, Ozturk N, Ozturk D, Okyar A. ABC transporters: circadian rhythms and sex-related differences. Journal of Marmara University Institute of Health Sciences. Published online 2013:1. doi:10.5455/musbed.20130306115105

41. Desprez F, Ung DC, Vourc’h P, Jeanne M, Laumonnier F. Contribution of the dihydropyrimidinase-like proteins family in synaptic physiology and in neurodevelopmental disorders. Front Neurosci. 2023;17. doi:10.3389/fnins.2023.1154446

42. Nousbeck J, Spiegel R, Ishida-Yamamoto A, et al. Alopecia, Neurological Defects, and Endocrinopathy Syndrome Caused by Decreased Expression of RBM28, a Nucleolar Protein Associated with Ribosome Biogenesis. The American Journal of Human Genetics. 2008;82(5):1114–1121. doi:10.1016/j.ajhg.2008.03.014

43. Laccone F, Schoner K, Krabichler B, et al. Desbuquois dysplasia type I and fetal hydrops due to novel mutations in the CANT1 gene. European Journal of Human Genetics. 2011;19(11):1133–1137. doi:10.1038/ejhg.2011.101

44. Robinson-Shelton A, Malow BA. Sleep Disturbances in Neurodevelopmental Disorders. Curr Psychiatry Rep. 2016;18(1):6. doi:10.1007/s11920-015-0638-1

45. Kamara D, Beauchaine TP. A Review of Sleep Disturbances among Infants and Children with Neurodevelopmental Disorders. Rev J Autism Dev Disord. 2020;7(3):278–294. doi:10.1007/s40489-019-00193-8

46. del Castillo-Izquierdo Á, Moreno-Navarrete JM, Latorre J, et al. DPP9 as a Potential Novel Mediator in Gastrointestinal Virus Infection. Antioxidants. 2022;11(11):2177. doi:10.3390/antiox11112177

47. Zhao YB, Wang SZ, Guo WT, et al. Hippocampal dipeptidyl peptidase 9 bidirectionally regulates memory associated with synaptic plasticity. J Adv Res. Published online October 2024. doi:10.1016/j.jare.2024.09.031

48. Guan FHX, Bailey CG, Metierre C, et al. The antiproliferative ELF2 isoform, ELF2B, induces apoptosis in vitro and perturbs early lymphocytic development in vivo. J Hematol Oncol. 2017;10(1):75. doi:10.1186/s13045-017-0446-7

49. Ahmad H, Requena T, Frejo L, et al. Clinical and Functional Characterization of a Missense ELF2 Variant in a CANVAS Family. Front Genet. 2018;9. doi:10.3389/fgene.2018.00085

