## Supplementary Table for "Genetics of Sleepwalking: Insights from whole exome sequencing"

**Supplementary Table 1: Detailed clinical information of each individual in the SW cohort.**

**21-25, 26-30, 31-35, 36-40, 41-45, 46-50, 51-etc**

| Region | Gender | Assessment Age | Assessment Age Group | Age Onset Group | FamilyHx  FirstDegree | FamilyHx  Details | Severity | Comorbidity | Comorbidity classification | Episodes Frequency |
| --- | --- | --- | --- | --- | --- | --- | --- | --- | --- | --- |
| Montreal | Female | 46-50 | Adult | Adult | Positive | mother, 2 brothers | Severe | no |  | Frequent |
| Montreal | Female | 41-45 | Adult | Child | Negative |  | Moderate | yes | Insomnia | Infrequent |
| Montreal | Female | 31-35 | Adult | Adult | Negative |  | Moderate | no |  | Frequent |
| Montreal | Female | 21-25 | Adult | Teen | Positive | mother, sister | Moderate | no |  | Frequent |
| Montreal | Female | 21-25 | Adult | Child | Positive | mother | Mild | no |  | Infrequent |
| Montreal | Female | 36-40 | Adult | Adult | Positive | 2 daugthers | Moderate | no |  | Frequent |
| Montreal | Female | 26-30 | Adult | Child | Positive | mother, sister | Moderate | yes | Insomnia | Infrequent |
| Montreal | Female | 31-35 | Adult | Child | Positive | sister | Moderate | no |  | Frequent |
| Montreal | Female | 26-30 | Adult | Child | Negative |  | Moderate | no |  | Frequent |
| Montreal | Female | 11-15 | Teen | Teen | Negative |  | Moderate | yes | Insomnia | Frequent |
| Montreal | Female | 31-35 | Adult | Child | Positive | brother | Severe | yes | Insomnia | Frequent |
| Montreal | Female | 26-30 | Adult | Teen | Negative |  | Moderate | no |  | Frequent |
| Montreal | Female | 36-40 | Adult | Child | Negative |  | Moderate | no |  | Frequent |
| Montreal | Female | 46-50 | Adult | Adult | Negative |  | Severe | no |  | Infrequent |
| Montreal | Female | 26-20 | Adult | Child | Positive | mother, brother | Moderate | no |  | Frequent |
| Montreal | Female | 51-55 | Adult | Child | Positive | daugther | Moderate | yes | Sleep Apnea | Infrequent |
| Montreal | Female | 41-45 | Adult | Adult | Positive | daugther | Severe | no |  | Frequent |
| Montreal | Female | 26-30 | Adult | Teen | Negative |  | Moderate | no |  | Frequent |
| Montreal | Female | 21-25 | Adult | Teen | Positive | father, brother | Moderate | no |  | Frequent |
| Montreal | Female | 21-25 | Adult | Teen | Positive | brother | Moderate | no |  | Frequent |
| Montreal | Female | 16-20 | Teen | Child | Negative |  | Moderate | no |  | Frequent |
| Montreal | Female | 36-40 | Adult | Child | Negative |  | Moderate | no |  | Frequent |
| Montreal | Female | 21-25 | Adult | Child | Negative |  | Moderate | no |  | Frequent |
| Montreal | Female | 31-35 | Adult | Child | Positive | brother | Moderate | yes | Insomnia | Frequent |
| Montreal | Female | 11-15 | Teen | Teen | Positive | mother | Moderate | no |  | Infrequent |
| Montreal | Female | 41-45 | Adult | Child | Positive | sister | Moderate | no |  | Frequent |
| Montreal | Female | 21-25 | Adult | Teen | Negative |  | Moderate | no |  | Frequent |
| Montreal | Female | 36-40 | Adult | Teen | Negative |  | Moderate | no |  | Frequent |
| Montreal | Female | 21-25 | Adult | Adult | Positive | mother, sister | Severe | no |  | Frequent |
| Montreal | Female | 41-45 | Adult | Teen | Positive | brother | Severe | yes | Insomnia | Frequent |
| Montreal | Female | 41-45 | Adult | Child | NA |  | Moderate | yes | Sleep Apnea | NA |
| Montreal | Female | 56-60 | Adult | Adult | Positive | daugther | Moderate | yes | Sleep Apnea | Frequent |
| Montreal | Female | 41-45 | Adult | Child | NA |  | Moderate | yes | Insomnia | Frequent |
| Montreal | Female | 46-50 | Adult | Teen | Negative |  | Moderate | yes | Insomnia | Frequent |
| Montreal | Female | 21-25 | Adult | Child | NA |  | Moderate | no |  | Frequent |
| Montreal | Female | 56-60 | Adult | Child | Negative |  | Moderate | no |  | Frequent |
| Montreal | Female | 16-20 | Adult | Child | Negative |  | Moderate | no |  | Infrequent |
| Montreal | Female | 31-35 | Adult | Child | Negative |  | Moderate | no |  | Frequent |
| Montreal | Female | 26-30 | Adult | Child | Positive | father, sister | Moderate | no |  | Infrequent |
| Montreal | Female | 36-40 | Adult | Child | Negative |  | Moderate | no |  | Frequent |
| Montreal | Female | 31-35 | Adult | Child | Positive | father | Moderate | no |  | Infrequent |
| Montreal | Female | 31-35 | Adult | Teen | Positive | mother | Moderate | no |  | Frequent |
| Montreal | Female | 16-20 | Adult | Teen | Positive | 2 brothers | Moderate | no |  | Frequent |
| Montreal | Female | 21-25 | Adult | Child | Positive | sister | Moderate | no |  | Frequent |
| Montreal | Female | 41-45 | Adult | Child | Positive | mother, daugthers | Moderate | no |  | Infrequent |
| Montreal | Female | 36-40 | Adult | Child | Positive | father, 2 daugthers | Moderate | yes | Insomnia | Frequent |
| Montreal | Female | 26-30 | Adult | Child | Positive | mother, brother | Moderate | no |  | Frequent |
| Montreal | Female | 26-30 | Adult | Adult | Positive | sister | Moderate | no |  | Infrequent |
| Montreal | Female | 26-30 | Adult | Child | Negative |  | Moderate | no |  | Frequent |
| Montreal | Female | 36-40 | Adult | Child | Negative |  | Moderate | no |  | Infrequent |
| Montreal | Female | 31-35 | Adult | Teen | Positive | father, 2 brothers, 1 sister | Moderate | no |  | Frequent |
| Montreal | Female | 21-25 | Adult | Child | Positive | mother, father | Moderate | no |  | Frequent |
| Montreal | Female | 36-40 | Adult | Child | Negative |  | Moderate | no |  | Infrequent |
| Montreal | Female | 46-50 | Adult | Child | Negative |  | Moderate | yes | Sleep Apnea | Infrequent |
| Montreal | Female | 21-25 | Adult | Child | Positive | mother | Moderate | no |  | Frequent |
| Montreal | Female | 31-35 | Adult | Adult | Positive | father, sister | Moderate | no |  | Frequent |
| Montreal | Female | 21-25 | Adult | Adult | Positive | father | Moderate | no |  | Frequent |
| Montreal | Female | 31-35 | Adult | Child | Positive | father | Moderate | no |  | Infrequent |
| Montreal | Female | 21-25 | Adult | Teen | Positive | 2 sisters | Moderate | no |  | Frequent |
| Montreal | Female | 26-30 | Adult | Child | Positive | mother | Moderate | no |  | Frequent |
| Montreal | Female | 21-25 | Adult | Child | Negative |  | Moderate | yes | Insomnia | Frequent |
| Montreal | Female | 26-30 | Adult | Child | Negative |  | Moderate | no |  | NA |
| Montreal | Female | 51-55 | Adult | Child | NA |  | Moderate | yes | Sleep Apnea | NA |
| Montreal | Female | 46-50 | Adult | Child | Positive | mother, father | Moderate | no |  | Frequent |
| Montreal | Female | 36-40 | Adult | Child | Negative |  | Moderate | yes | Insomnia | Frequent |
| Montreal | Female | 31-35 | Adult | Child | Negative |  | Moderate | no |  | Frequent |
| Montreal | Female | 31-35 | Adult | Child | Negative |  | Moderate | no |  | Frequent |
| Montreal | Female | 46-50 | Adult | Child | Negative |  | Moderate | no |  | Frequent |
| Montreal | Female | 36-40 | Adult | Child | Negative |  | Moderate | yes | Insomnia | Infrequent |
| Montreal | Female | 26-30 | Adult | Adult | Negative |  | Severe | no |  | Frequent |
| Montreal | Female | 26-30 | Adult | Child | NA |  | Moderate | no |  | NA |
| Montreal | Female | 41-45 | Adult | Child | Positive | daughter, sons | Severe | no |  | Frequent |
| Montreal | Female | 36-40 | Adult | Child | Negative | niece | Moderate | no |  | Frequent |
| Montreal | Female | 26-30 | Adult | Child | Positive | mother, father | Moderate | no |  | Frequent |
| Montreal | Female | 21-25 | Adult | Child | Positive | father | Severe | no |  | Frequent |
| Montreal | Female | 31-35 | Adult | Child | Positive | mother | Moderate | no |  | Frequent |
| Montreal | Female | 36-40 | Adult | Child | Negative |  | Moderate | yes | Sleep Apnea | Frequent |
| Montreal | Male | 26-30 | Adult | Child | Negative |  | Moderate | yes | Sleep Apnea | Frequent |
| Montreal | Male | 31-35 | Adult | Child | Negative |  | Moderate | yes | Sleep Apnea | Frequent |
| Montreal | Male | 21-25 | Adult | Teen | Positive | father | Severe | no |  | Frequent |
| Montreal | Male | 61-65 | Adult | Adult | Negative |  | Moderate | yes | Sleep Apnea | NA |
| Montreal | Male | 31-35 | Adult | Teen | Positive | sons | Moderate | no |  | Frequent |
| Montreal | Male | 21-25 | Adult | Child | Negative |  | Moderate | no |  | Frequent |
| Montreal | Male | 36-40 | Adult | Child | Negative |  | Moderate | no |  | Frequent |
| Montreal | Male | 31-35 | Adult | Adult | Positive | daughter | Moderate | no |  | Frequent |
| Montreal | Male | 36-40 | Adult | Adult | Negative |  | Moderate | no |  | Frequent |
| Montreal | Male | 31-35 | Adult | Child | Negative |  | Moderate | yes | Sleep Apnea | Frequent |
| Montreal | Male | 46-50 | Adult | Teen | Negative |  | Moderate | yes | Sleep Apnea | Infrequent |
| Montreal | Male | 36-40 | Adult | Teen | Positive | mother | Moderate | yes | Insomnia | Frequent |
| Montreal | Male | 21-25 | Adult | Child | Negative |  | Moderate | no |  | Infrequent |
| Montreal | Male | 21-25 | Adult | Teen | Negative |  | Moderate | no |  | Frequent |
| Montreal | Male | 56-60 | Adult | Child | Positive | father | Moderate | no |  | Frequent |
| Montreal | Male | 26-30 | Adult | Child | Positive | mother | Moderate | no |  | Frequent |
| Montreal | Male | 66-70 | Adult | Adult | NA |  | Moderate | no |  | NA |
| Montreal | Male | 56-60 | Adult | Adult | Negative |  | Moderate | yes | Insomnia | Frequent |
| Montreal | Male | 61-65 | Adult | Child | Negative |  | Moderate | yes | Insomnia | Frequent |
| Montreal | Male | 46-50 | Adult | Adult | Positive | brother | Moderate | no |  | NA |
| Montreal | Male | 16-20 | Adult | Child | Positive | father | Moderate | no |  | Frequent |
| Montreal | Male | 61-65 | Adult | Teen | Positive |  | Moderate | no |  | Infrequent |
| Montreal | Male | 26-30 | Adult | Child | Positive | father | Moderate | no |  | Infrequent |
| Montreal | Male | 51-55 | Adult | Teen | Positive | mother, brother, sons | Moderate | no |  | Frequent |
| Montreal | Male | 31-35 | Adult | Adult | Positive | sons | Severe | no |  | Frequent |
| Montreal | Male | 26-30 | Adult | Child | Negative |  | Moderate | no |  | Frequent |
| Montreal | Male | 41-45 | Adult | Adult | Negative |  | Severe | no |  | Frequent |
| Montreal | Male | 36-40 | Adult | Child | Positive | father, daugther | Moderate | no |  | Frequent |
| Montreal | Male | 41-45 | Adult | Child | NA |  | Moderate | no |  | Frequent |
| Montreal | Male | 66-70 | Adult | Adult | Negative |  | Moderate | yes | Sleep Apnea | Frequent |
| Montreal | Male | 31-35 | Adult | Child | Negative |  | Moderate | no |  | Frequent |
| Montreal | Male | 36-40 | Adult | Teen | Positive | mother | Moderate | yes | Sleep Apnea | Frequent |
| Montreal | Male | 16-20 | Adult | Teen | Negative |  | Severe | no |  | Infrequent |
| Montreal | Male | 31-35 | Adult | Child | Positive |  | Severe | no |  | Frequent |
| Montreal | Male | 46-50 | Adult | Child | Negative |  | Moderate | no |  | Frequent |
| Montreal | Male | 21-25 | Adult | Child | Negative |  | Moderate | no |  | Infrequent |
| Montreal | Male | 31-35 | Adult | Adult | Negative |  | Moderate | yes | Sleep Apnea | Frequent |
| Montreal | Male | 36-40 | Adult | Child | Positive | father, daugther | Moderate | no |  | Frequent |
| Montreal | Male | 31-35 | Adult | Child | Negative |  | Moderate | no |  | Frequent |
| Montreal | Male | 36-40 | Adult | Child | Negative |  | Moderate | yes | Insomnia | Frequent |
| Montreal | Male | 36-40 | Adult | Child | Negative |  | Moderate | no |  | Frequent |
| Montreal | Male | 41-45 | Adult | Child | Positive |  | Moderate | yes | Sleep Apnea | Infrequent |
| Montreal | Male | 21-25 | Adult | Child | Positive | father, brothers | Moderate | no |  | Frequent |
| Montreal | Male | 31-35 | Adult | Child | Positive | daughter | Moderate | no |  | Frequent |
| Montreal | Male | 26-30 | Adult | Child | Negative |  | Moderate | no |  | Frequent |
| Montreal | Male | 36-40 | Adult | Child | NA |  | Moderate | yes | Insomnia | NA |
| Montreal | Male | 31-35 | Adult | Child | Negative |  | Moderate | no |  | Frequent |
| Montreal | Male | 21-25 | Adult | Child | Negative |  | Moderate | no |  | Frequent |
| Montreal | Male | 26-30 | Adult | Child | Negative |  | Moderate | yes | Insomnia | Frequent |
| Montreal | Male | 26-30 | Adult | Child | NA |  | Moderate | no |  | Frequent |
| Montreal | Male | 26-30 | Adult | Child | Positive | mother | Moderate | no |  | Frequent |
| Montreal | Male | 31-35 | Adult | Child | Positive | father, mother, sister | Severe | no |  | Frequent |
| France | Female | 11-15 | Child | Child | Positive | parents, siblings, or children | Moderate | no |  | Frequent |
| France | Female | 11-15 | Teen | Teen | Positive | parents, siblings, or children | Moderate | no |  | Frequent |
| France | Female | 21-25 | Adult | Child | Negative |  | Moderate | no |  | Frequent |
| France | Female | 21-25 | Adult | Teen | Negative |  | Moderate | no |  | Frequent |
| France | Female | 36-40 | Adult | Teen | Positive | parents, siblings, or children | Moderate | no |  | Frequent |
| France | Female | 26-30 | Adult | Child | Negative |  | Moderate | no |  | Frequent |
| France | Female | 21-25 | Adult | Child | NA |  | Moderate | no |  | NA |
| France | Female | 16-20 | Adult | Teen | Negative |  | Moderate | no |  | Frequent |
| France | Female | 36-40 | Adult | Child | Positive | parents, siblings, or children | Moderate | no |  | Frequent |
| France | Female | 6-10 | Child | Child | Positive | parents, siblings, or children | Moderate | no |  | NA |
| France | Female | 26-30 | Adult | Child | Positive | parents, siblings, or children | Moderate | no |  | Frequent |
| France | Female | 21-25 | Adult | Child | Positive | parents, siblings, or children | Moderate | no |  | Frequent |
| France | Female | 51-55 | Adult | Adult | Negative |  | Moderate | no |  | Frequent |
| France | Female | 21-25 | Adult | Teen | Negative |  | Moderate | no |  | Frequent |
| France | Female | 41-45 | Adult | Child | Positive | parents, siblings, or children | Moderate | yes | RLS | Frequent |
| France | Female | 56-60 | Adult | Child | Positive | parents, siblings, or children | Moderate | no |  | Frequent |
| France | Female | 26-30 | Adult | Child | Negative |  | Moderate | yes | RLS | Frequent |
| France | Female | 41-45 | Adult | Child | Positive | parents, siblings, or children | Moderate | yes | RLS | Frequent |
| France | Female | 16-20 | Adult | Teen | Positive | parents, siblings, or children | Moderate | no |  | Frequent |
| France | Female | 21-25 | Adult | Child | Negative |  | Moderate | no |  | Frequent |
| France | Female | 21-25 | Adult | Child | Negative |  | Moderate | no |  | Frequent |
| France | Female | 21-25 | Adult | Child | Positive | parents, siblings, or children | Moderate | no |  | Frequent |
| France | Female | 21-25 | Adult | Adult | Negative |  | Moderate | no |  | Frequent |
| France | Female | 26-30 | Adult | Child | Negative |  | Moderate | no |  | Frequent |
| France | Female | 36-40 | Adult | Child | Positive | parents, siblings, or children | Moderate | no |  | Frequent |
| France | Female | 26-30 | Adult | Child | NA |  | Moderate | no |  | NA |
| France | Female | 31-35 | Adult | Child | Positive | parents, siblings, or children | Moderate | no |  | Frequent |
| France | Female | 6-10 | Child | Child | Positive | parents, siblings, or children | Moderate | no |  | Frequent |
| France | Female | 21-25 | Adult | Child | Positive | parents, siblings, or children | Moderate | no |  | Frequent |
| France | Female | 26-30 | Adult | Child | Negative |  | Moderate | no |  | Frequent |
| France | Female | 31-35 | Adult | Child | Positive | parents, siblings, or children | Moderate | no |  | Frequent |
| France | Female | 26-30 | Adult | Adult | Negative |  | Moderate | no |  | Frequent |
| France | Female | 31-35 | Adult | Child | Positive | parents, siblings, or children | Moderate | no |  | Frequent |
| France | Female | 21-25 | Adult | Child | Negative |  | Moderate | no |  | Frequent |
| France | Female | 21-25 | Adult | Teen | Negative |  | Moderate | no |  | Frequent |
| France | Female | 31-35 | Adult | Child | Negative |  | Moderate | no |  | Frequent |
| France | Female | 26-30 | Adult | Child | Negative |  | Moderate | no |  | Frequent |
| France | Female | 26-30 | Adult | Child | Positive | parents, siblings, or children | Moderate | no |  | Frequent |
| France | Female | 21-25 | Adult | Child | Negative |  | Moderate | no |  | Frequent |
| France | Female | 16-20 | Adult | Child | Negative |  | Moderate | no |  | Frequent |
| France | Female | 21-25 | Adult | Child | Negative |  | Moderate | no |  | Frequent |
| France | Female | 31-35 | Adult | Child | Positive | parents, siblings, or children | Moderate | no |  | Frequent |
| France | Female | 21-25 | Adult | Child | Negative |  | Moderate | no |  | Frequent |
| France | Female | 21-25 | Adult | Child | Positive | parents, siblings, or children | Moderate | no |  | Frequent |
| France | Female | 31-35 | Adult | Child | Positive | parents, siblings, or children | Moderate | no |  | Frequent |
| France | Female | 31-35 | Adult | Child | Positive | parents, siblings, or children | Moderate | no |  | Frequent |
| France | Female | 21-25 | Adult | Child | Negative |  | Moderate | no |  | Frequent |
| France | Female | 31-35 | Adult | Child | Negative |  | Moderate | no |  | Frequent |
| France | Female | 21-25 | Adult | Child | Negative |  | Moderate | no |  | Frequent |
| France | Female | 26-30 | Adult | Adult | Negative |  | Moderate | no |  | Frequent |
| France | Female | 21-25 | Adult | Child | Positive | parents, siblings, or children | Moderate | no |  | Frequent |
| France | Female | 46-50 | Adult | Child | Positive | parents, siblings, or children | Moderate | no |  | Frequent |
| France | Female | 26-30 | Adult | Teen | Negative |  | Moderate | no |  | NA |
| France | Female | 51-55 | Adult | Child | Positive | parents, siblings, or children | Moderate | no |  | Frequent |
| France | Female | 16-20 | Adult | Child | Negative |  | Moderate | no |  | Frequent |
| France | Female | 46-50 | Adult | Child | Positive | parents, siblings, or children | Moderate | no |  | Frequent |
| France | Female | 21-25 | Adult | Teen | Positive | parents, siblings, or children | Moderate | no |  | Frequent |
| France | Female | 36-40 | Adult | Child | Negative |  | Moderate | no |  | Frequent |
| France | Female | 31-35 | Adult | Child | Negative |  | Moderate | no |  | Frequent |
| France | Female | 46-50 | Adult | Child | Positive | parents, siblings, or children | Moderate | no |  | Frequent |
| France | Female | 26-30 | Adult | Child | Positive | parents, siblings, or children | Moderate | no |  | Frequent |
| France | Female | 16-20 | Adult | Child | Negative |  | Moderate | no |  | Frequent |
| France | Male | 21-25 | Adult | Child | Positive | parents, siblings, or children | Moderate | no |  | Frequent |
| France | Male | 21-25 | Adult | Child | Positive | parents, siblings, or children | Moderate | no |  | Frequent |
| France | Male | 31-35 | Adult | Child | Positive | parents, siblings, or children | Moderate | no |  | NA |
| France | Male | 16-20 | Adult | Child | Positive | parents, siblings, or children | Moderate | no |  | Frequent |
| France | Male | 6-10 | Child | Child | Positive | parents, siblings, or children | Moderate | no |  | Frequent |
| France | Male | 16-20 | Teen | Child | Positive | parents, siblings, or children | Moderate | no |  | Frequent |
| France | Male | 6-10 | Child | Child | Positive | parents, siblings, or children | Moderate | no |  | Frequent |
| France | Male | 51-55 | Adult | Child | Positive | parents, siblings, or children | Moderate | no |  | Frequent |
| France | Male | 21-25 | Adult | Teen | Negative |  | Moderate | no |  | Frequent |
| France | Male | 26-30 | Adult | Adult | NA |  | Moderate | no |  | NA |
| France | Male | 11-15 | Child | Child | Positive | parents, siblings, or children | Moderate | no |  | Frequent |
| France | Male | 11-15 | Teen | Child | Positive | parents, siblings, or children | Moderate | no |  | Frequent |
| France | Male | 26-30 | Adult | Teen | Negative |  | Moderate | no |  | Frequent |
| France | Male | 16-20 | Teen | Child | Positive | parents, siblings, or children | Moderate | no |  | Frequent |
| France | Male | 16-20 | Teen | Teen | Positive | parents, siblings, or children | Moderate | no |  | Frequent |
| France | Male | 6-10 | Child | Child | Negative |  | Moderate | no |  | Frequent |
| France | Male | 11-15 | Teen | Child | Negative |  | Moderate | no |  | Frequent |
| France | Male | 6-10 | Child | Child | Positive | parents, siblings, or children | Moderate | no |  | NA |
| France | Male | 11-15 | Teen | Child | Negative |  | Moderate | no |  | Frequent |
| France | Male | 31-35 | Adult | Child | Positive | parents, siblings, or children | Moderate | yes | RLS | Frequent |
| France | Male | 36-40 | Adult | Teen | Negative |  | Severe | no |  | Frequent |
| France | Male | 41-45 | Adult | Adult | Negative |  | Moderate | no |  | Frequent |
| France | Male | 26-30 | Adult | Child | Negative |  | Moderate | no |  | Frequent |
| France | Male | 36-40 | Adult | Adult | Negative |  | Moderate | no |  | Frequent |
| France | Male | 36-40 | Adult | Child | Positive | parents, siblings, or children | Moderate | no |  | Frequent |
| France | Male | 26-30 | Adult | Child | Positive | parents, siblings, or children | Moderate | no |  | Frequent |
| France | Male | 46-50 | Adult | Child | Positive | parents, siblings, or children | Moderate | no |  | Frequent |
| France | Male | 11-15 | Teen | Child | NA |  | Moderate | no |  | Frequent |
| France | Male | 6-10 | Child | Child | Negative |  | Moderate | no |  | Frequent |
| France | Male | 41-45 | Adult | Child | Positive | parents, siblings, or children | Moderate | no |  | Frequent |
| France | Male | 31-35 | Adult | Child | NA |  | Moderate | no |  | Frequent |
| France | Male | 31-35 | Adult | Adult | Negative |  | Moderate | yes | RLS | Frequent |
| France | Male | 21-25 | Adult | Child | Positive | parents, siblings, or children | Moderate | yes | RLS | Infrequent |
| France | Male | 16-20 | Teen | Child | Positive | parents, siblings, or children | Moderate | no |  | Frequent |
| France | Male | 11-15 | Teen | Child | Positive | parents, siblings, or children | Moderate | no |  | Frequent |
| France | Male | 21-25 | Adult | Teen | Negative |  | Moderate | no |  | Frequent |
| France | Male | 16-20 | Adult | Child | Negative |  | Moderate | no |  | Frequent |
| France | Male | 41-45 | Adult | Teen | Positive | parents, siblings, or children | Moderate | no |  | Frequent |
| France | Male | 11-15 | Teen | Child | Positive | parents, siblings, or children | Moderate | no |  | Frequent |
| France | Male | 26-30 | Adult | Child | Negative |  | Moderate | no |  | Frequent |
| France | Male | 26-30 | Adult | Teen | Positive | parents, siblings, or children | Moderate | no |  | Frequent |
| France | Male | 31-35 | Adult | Child | Negative |  | Moderate | no |  | Frequent |
| France | Male | 21-25 | Adult | Child | Positive | parents, siblings, or children | Moderate | no |  | Frequent |
| France | Male | 21-25 | Adult | Child | Negative |  | Moderate | no |  | Frequent |
| France | Male | 31-35 | Adult | Child | Positive | parents, siblings, or children | Moderate | yes | RLS | Frequent |
| France | Male | 21-25 | Adult | Teen | Negative |  | Moderate | no |  | Frequent |
| France | Male | 26-30 | Adult | Teen | Positive | parents, siblings, or children | Moderate | no |  | Frequent |
| France | Male | 31-35 | Adult | Teen | Negative |  | Moderate | no |  | Frequent |
| France | Male | 31-35 | Adult | Child | Positive | parents, siblings, or children | Moderate | yes | RLS | Frequent |
| France | Male | 31-35 | Adult | Child | Negative |  | Moderate | no |  | Frequent |
| France | Male | 41-45 | Adult | Child | Positive | parents, siblings, or children | Moderate | no |  | Frequent |
| France | Male | 6-10 | Child | Child | Positive | parents, siblings, or children | Moderate | no |  | Frequent |
| France | Male | 11-15 | Teen | Child | Positive | parents, siblings, or children | Moderate | no |  | Frequent |
| France | Male | 31-35 | Adult | Child | Negative |  | Moderate | no |  | Frequent |
| France | Male | 31-35 | Adult | Child | Negative |  | Moderate | no |  | Frequent |
| France | Male | 31-35 | Adult | Child | Positive | parents, siblings, or children | Moderate | no |  | Frequent |
| France | Male | 36-40 | Adult | Child | Negative |  | Severe | yes | RLS | Frequent |
| France | Male | 26-30 | Adult | Child | Negative |  | Moderate | no |  | Frequent |
| France | Male | 21-25 | Adult | Child | Negative |  | Moderate | no |  | Frequent |
| France | Male | 16-20 | Adult | Adult | Negative |  | Moderate | no |  | Frequent |
| France | Male | 6-10 | Child | Child | Positive | parents, siblings, or children | Moderate | no |  | Frequent |
| France | Male | 26-30 | Adult | Child | Positive | parents, siblings, or children | Moderate | no |  | Frequent |
| France | Male | 26-30 | Adult | Child | Positive | parents, siblings, or children | Moderate | no |  | Frequent |

**Supplementary Table 2: Detailed clinical information of each individual in the Control cohort.**

| **Region** | **Gender** | **Assessment Age** | **Assessment Age Group** | **Comorbidity** |
| --- | --- | --- | --- | --- |
| Montreal | Female | > 18 | Adult | RLS |
| Montreal | Female | > 18 | Adult | Insomnia |
| Montreal | Female | > 18 | Adult | Insomnia |
| Montreal | Male | > 18 | Adult | Insomnia |
| Montreal | Female | > 18 | Adult | Insomnia |
| Montreal | Female | > 18 | Adult | Insomnia |
| Montreal | Female | > 18 | Adult | Insomnia |
| Montreal | Female | > 18 | Adult | Insomnia |
| Montreal | Female | > 18 | Adult | Insomnia |
| Montreal | Female | > 18 | Adult | Insomnia |
| Montreal | Male | > 18 | Adult | Insomnia |
| Montreal | Male | > 18 | Adult | Sleep Apnea |
| Montreal | Female | > 18 | Adult | Insomnia |
| Montreal | Male | > 18 | Adult |  |
| Montreal | Male | > 18 | Adult | Insomnia |
| Montreal | Female | > 18 | Adult | Insomnia |
| Montreal | Female | > 18 | Adult | RLS |
| Montreal | Male | > 18 | Adult | Sleep Apnea |
| Montreal | Female | > 18 | Adult | Insomnia |
| Montreal | Male | > 18 | Adult | Insomnia |
| Montreal | Female | > 18 | Adult | Insomnia |
| Montreal | Male | > 18 | Adult | RLS |
| Montreal | Female | > 18 | Adult | Insomnia |
| Montreal | Female | > 18 | Adult | Insomnia |
| Montreal | Female | > 18 | Adult | Insomnia |
| Montreal | Male | > 18 | Adult | Insomnia |
| Montreal | Female | > 18 | Adult | Insomnia |
| Montreal | Female | > 18 | Adult | RLS |
| Montreal | Male | > 18 | Adult | Insomnia |
| Montreal | Female | > 18 | Adult | Insomnia |
| Montreal | Male | > 18 | Adult | RLS |
| Montreal | Male | > 18 | Adult | Insomnia |
| Montreal | Male | > 18 | Adult | Insomnia |
| Montreal | Male | > 18 | Adult | Sleep Apnea |
| Montreal | Male | > 18 | Adult | Insomnia |
| Montreal | Female | > 18 | Adult | Insomnia |
| Montreal | Female | > 18 | Adult | Insomnia |
| Montreal | Female | > 18 | Adult | Insomnia |
| Montreal | Male | > 18 | Adult | RLS |
| Montreal | Male | > 18 | Adult | Insomnia |
| Montreal | Female | > 18 | Adult | Insomnia |
| Montreal | Female | > 18 | Adult | Insomnia |
| Montreal | Male | > 18 | Adult | Insomnia |
| Montreal | Male | > 18 | Adult | Insomnia |
| Montreal | Male | > 18 | Adult | Sleep Apnea |
| Montreal | Female | > 18 | Adult | Insomnia |
| Montreal | Male | > 18 | Adult | Insomnia |
| Montreal | Male | > 18 | Adult | RLS |
| Montreal | Male | > 18 | Adult | RLS |
| Montreal | Female | > 18 | Adult | Sleep Apnea |
| Montreal | Female | > 18 | Adult | Insomnia |
| Montreal | Male | > 18 | Adult | Insomnia |
| Montreal | Female | > 18 | Adult | RLS |
| Montreal | Female | > 18 | Adult | Insomnia |
| Montreal | Male | > 18 | Adult | Insomnia |
| Montreal | Female | > 18 | Adult | Insomnia |
| Montreal | Female | > 18 | Adult | Insomnia |
| Montreal | Male | > 18 | Adult | Insomnia |
| Montreal | Female | > 18 | Adult | Insomnia |
| Montreal | Female | > 18 | Adult | Insomnia |
| Montreal | Female | > 18 | Adult | RLS |
| Montreal | Female | > 18 | Adult | Insomnia |
| Montreal | Male | > 18 | Adult | RLS |
| Montreal | Male | > 18 | Adult | Sleep Apnea |
| France | Female | 21-25 | Adult |  |
| France | Female | 26-30 | Adult | Insomnia |
| France | Male | 46-50 | Adult |  |
| France | Female | 46-50 | Adult | Insomnia |
| France | Female | 46-50 | Adult | Autoimmune disease |
| France | Female | 21-25 | Adult |  |
| France | Female | 46-50 | Adult | RLS, Insomnia |
| France | Female | 36-40 | Adult | Insomnia |
| France | Female | 20 | Adult |  |
| France | Male | 36-40 | Adult |  |
| France | Female | 46-50 | Adult |  |
| France | Male | 51-55 | Adult |  |
| France | Female | 31-35 | Adult | Autoimmune disease, Insomnia |
| France | Female | 46-50 | Adult | Autoimmune disease |
| France | Female | 56-60 | Adult |  |
| France | Male | - | Adult |  |
| France | Male | 41-45 | Adult |  |
| France | Female | 46-50 | Adult | Insomnia |
| France | Male | 31-35 | Adult | Insomnia |
| France | Male | 36-40 | Adult |  |
| France | Female | 31-35 | Adult |  |
| France | Male | 26-30 | Adult | Insomnia |
| France | Female | 51-55 | Adult | Autoimmune disease, Insomnia |
| France | Male | 51-55 | Adult |  |
| France | Male | 21-25 | Adult |  |
| France | Male | 26-30 | Adult |  |
| France | Male | 21-25 | Adult |  |
| France | Female | 31-35 | Adult |  |
| France | Female | 36-40 | Adult | Insomnia |
| France | Male | 31-35 | Adult |  |
| France | Female | 31-35 | Adult |  |
| France | Male | 26-30 | Adult |  |
| France | Female | 31-35 | Adult |  |
| France | Male | 26-30 | Adult |  |
| France | Female | 46-50 | Adult | Insomnia |
| France | Male | 61-65 | Adult | Autoimmune disease |
| France | Male | 26-30 | Adult |  |
| France | Female | 21-25 | Adult |  |
| France | Female | 56-60 | Adult |  |
| France | Female | 36-40 | Adult |  |
| France | Female | 46-50 | Adult |  |
| France | Male | 21-25 | Adult |  |
| France | Male | 21-25 | Adult |  |
| France | Female | 21-25 | Adult |  |
| France | Female | 21-25 | Adult |  |
| France | Female | 21-25 | Adult |  |
| France | Male | 21-25 | Adult |  |
| France | Female | 61-65 | Adult | Insomnia |
| France | Male | 61-65 | Adult |  |
| France | Male | 21-25 | Adult | Insomnia |
| France | Female | 21-25 | Adult |  |
| France | Male | 16-20 | Adult | Insomnia |
| France | Male | 21-25 | Adult | Insomnia |
| France | Male | 21-25 | Adult | Autoimmune disease |
| France | Male | 21-25 | Adult | Insomnia |
| France | Male | 51-55 | Adult | Insomnia |
| France | Male | 31-35 | Adult |  |
| France | Male | 36-40 | Adult | Insomnia |
| France | Male | 31-35 | Adult | Autoimmune disease |
| France | Male | 16-20 | Adult |  |

**Supplementary Table 3:** **Fisher’s Exact Test Results for SW Case-Control**. Comparison. The table shows the 99 genes significantly enriched (p < 0.05) with the Odds Ratios and 95% Confidence Intervals.

| **Genes** | **p value** | **odd ratio** | **Confidence Intervals (CI) lowerbound** | **Confidence Intervals (CI) highbound** |
| --- | --- | --- | --- | --- |
| AAR2 | 0.04 | inf | 0.45 | 137.63 |
| ABCC10 | 0.03 | 3.28 | 0.97 | 11.14 |
| ACOT12 | 0.04 | inf | 0.45 | 137.63 |
| ADAMTS19 | 0.02 | inf | 0.57 | 169.17 |
| ADGRL2 | 0.03 | 4.18 | 0.96 | 18.25 |
| AIFM2 | 0.03 | inf | 0.51 | 153.40 |
| ANOS1 | 0.03 | inf | 0.51 | 153.40 |
| APC2 | 0.02 | 6.89 | 0.90 | 52.69 |
| APLF | 0.02 | 6.89 | 0.90 | 52.69 |
| ART5 | 0.03 | inf | 0.51 | 153.40 |
| BABAM2 | 0.03 | inf | 0.51 | 153.40 |
| BICDL2 | 0.04 | 5.90 | 0.76 | 45.67 |
| CALML5 | 0.04 | inf | 0.45 | 137.63 |
| CAMKK2 | 0.03 | inf | 0.51 | 153.40 |
| CANT1 | 0.01 | inf | 0.76 | 216.51 |
| CARNS1 | 0.02 | 6.89 | 0.90 | 52.69 |
| CEPT1 | 0.04 | inf | 0.45 | 137.63 |
| CGNL1 | 0.04 | 3.12 | 0.91 | 10.63 |
| CNGB3 | 0.03 | 6.40 | 0.83 | 49.18 |
| CNTD2 | 0.04 | inf | 0.45 | 137.63 |
| COL4A4 | 0.04 | inf | 0.45 | 137.63 |
| COQ3 | 0.04 | inf | 0.45 | 137.63 |
| CRAT | 0.02 | inf | 0.57 | 169.17 |
| CSF3R | 0.04 | inf | 0.45 | 137.63 |
| DCBLD2 | 0.02 | inf | 0.57 | 169.17 |
| DENND3 | 0.01 | inf | 0.63 | 184.95 |
| DIRC1 | 0.01 | inf | 0.63 | 184.95 |
| DNAH10 | 0.04 | 1.74 | 0.96 | 3.15 |
| DPP9 | 0.01 | inf | 0.76 | 216.51 |
| DPYSL5 | 0.04 | 5.90 | 0.76 | 45.67 |
| ECM2 | 0.04 | inf | 0.45 | 137.63 |
| ELF2 | 0.02 | 6.89 | 0.90 | 52.69 |
| ENPP1 | 0.01 | inf | 0.63 | 184.95 |
| ENPP3 | 0.02 | inf | 0.57 | 169.17 |
| ERCC4 | 0.05 | 3.69 | 0.84 | 16.26 |
| FRMD3 | 0.02 | inf | 0.57 | 169.17 |
| GLIS2 | 0.04 | inf | 0.45 | 137.63 |
| GNAT2 | 0.03 | inf | 0.51 | 153.40 |
| GPR132 | 0.04 | inf | 0.45 | 137.63 |
| HCRTR2 | 0.02 | inf | 0.57 | 169.17 |
| HEPH | 0.02 | inf | 0.57 | 169.17 |
| HGSNAT | 0.04 | inf | 0.45 | 137.63 |
| IDH2 | 0.04 | inf | 0.45 | 137.63 |
| IFI44L | 0.01 | inf | 0.63 | 184.95 |
| IGDCC4 | 0.04 | 5.90 | 0.76 | 45.67 |
| IPO4 | 0.05 | 3.69 | 0.84 | 16.26 |
| KCNH8 | 0.04 | inf | 0.45 | 137.63 |
| KIAA1109 | 0.04 | 3.12 | 0.91 | 10.63 |
| KIF7 | 0.03 | 4.18 | 0.96 | 18.25 |
| KRT12 | 0.04 | inf | 0.45 | 137.63 |
| KRT83 | 0.01 | inf | 0.63 | 184.95 |
| KRTAP12-2 | 0.03 | inf | 0.51 | 153.40 |
| MAGEC2 | 0.03 | inf | 0.51 | 153.40 |
| MCF2L2 | 0.02 | 7.38 | 0.97 | 56.20 |
| MFSD12 | 0.03 | 6.40 | 0.83 | 49.18 |
| MINK1 | 0.03 | inf | 0.51 | 153.40 |
| MOCS2 | 0.04 | 5.90 | 0.76 | 45.67 |
| MS4A18 | 0.01 | inf | 0.63 | 184.95 |
| NF1 | 0.02 | inf | 0.57 | 169.17 |
| NID1 | 0.04 | 5.90 | 0.76 | 45.67 |
| NOD2 | 0.03 | 2.95 | 1.01 | 8.60 |
| NPIPB13 | 0.00 | 9.10 | 2.18 | 38.08 |
| NUP155 | 0.04 | inf | 0.45 | 137.63 |
| PCSK4 | 0.03 | 6.40 | 0.83 | 49.18 |
| PHC1 | 0.04 | inf | 0.45 | 137.63 |
| PHGDH | 0.04 | inf | 0.45 | 137.63 |
| PNPLA2 | 0.02 | inf | 0.57 | 169.17 |
| PROB1 | 0.04 | inf | 0.45 | 137.63 |
| PTPRD | 0.04 | 5.90 | 0.76 | 45.67 |
| RASSF1 | 0.02 | inf | 0.57 | 169.17 |
| RBL2 | 0.04 | 5.90 | 0.76 | 45.67 |
| RBM28 | 0.03 | 6.40 | 0.83 | 49.18 |
| RHBDF1 | 0.02 | inf | 0.57 | 169.17 |
| RPL3L | 0.03 | inf | 0.51 | 153.40 |
| SEMA6B | 0.03 | 4.18 | 0.96 | 18.25 |
| SIRT1 | 0.03 | 3.28 | 0.97 | 11.14 |
| SLC25A30 | 0.04 | inf | 0.45 | 137.63 |
| SLC34A3 | 0.02 | inf | 0.57 | 169.17 |
| SLC3A1 | 0.04 | 3.94 | 0.90 | 17.26 |
| SLC9A8 | 0.02 | inf | 0.57 | 169.17 |
| SNTG2 | 0.04 | inf | 0.45 | 137.63 |
| SPNS2 | 0.03 | inf | 0.51 | 153.40 |
| SRRM2 | 0.02 | 2.39 | 1.04 | 5.47 |
| ST18 | 0.04 | inf | 0.45 | 137.63 |
| STARD8 | 0.03 | inf | 0.51 | 153.40 |
| SV2C | 0.03 | inf | 0.51 | 153.40 |
| TLR3 | 0.02 | inf | 0.57 | 169.17 |
| TMEM175 | 0.03 | 6.40 | 0.83 | 49.18 |
| TRIM32 | 0.03 | inf | 0.51 | 153.40 |
| TRPM5 | 0.01 | inf | 0.63 | 184.95 |
| TSR1 | 0.04 | 5.90 | 0.76 | 45.67 |
| UBASH3B | 0.03 | inf | 0.51 | 153.40 |
| USP17L13 | 0.01 | inf | 0.69 | 200.73 |
| WDR35 | 0.03 | inf | 0.51 | 153.40 |
| WWC2 | 0.04 | inf | 0.45 | 137.63 |
| ZFHX4 | 0.03 | 4.18 | 0.96 | 18.25 |
| ZFP91 | 0.01 | inf | 0.69 | 200.73 |
| ZNF41 | 0.03 | inf | 0.51 | 153.40 |
| ZSCAN10 | 0.01 | 5.41 | 1.26 | 23.20 |

*inf: infinity

**Supplementary Table 4: Contribution of Enriched Genes in the SW cohort.**

The table shows the number of counts of the significantly enriched genes in the SW cohort compared to the Control cohort. For each gene/variant the table includes the corresponding contribution scores and brain expression level.

| **Gene** | **Position** | **rsID** | **TotalCount Ctrl** | **TotalCount SW** | **Contribution score per allele (%)** | | **Contribution score per gene (%)** | **Contribution score (% in population)** | **Brain expression** | **Contribution to familial cases (%)** |
| --- | --- | --- | --- | --- | --- | --- | --- | --- | --- | --- |
| **NPIPB13** | chr16:30236217 | rs796810494 | 0 | 32 | 6.35 | 6.55 | | 13.10 | yes | 27% |
| **NPIPB13** | chr16:30237003 | rs1240720619 | 0 | 1 | 0.2 |  | |  |  |  |
| **DNAH10** | chr12:124270348 | rs147774367 | 0 | 2 | 0.4 | 4.59 | | 9.18 | no |  |
| **DNAH10** | chr12:124297903 | rs118146260 | 0 | 2 | 0.4 |  | |  |  |  |
| **DNAH10** | chr12:124317737 | rs199754209 | 0 | 2 | 0.4 |  | |  |  |  |
| **DNAH10** | chr12:124343814 | rs200692078 | 0 | 3 | 0.6 |  | |  |  |  |
| **DNAH10** | chr12:124362332 | rs186076361 | 1 | 5 | 0.99 |  | |  |  |  |
| **DNAH10** | chr12:124377896 | rs185566978 | 0 | 2 | 0.4 |  | |  |  |  |
| **DNAH10** | chr12:124409693 | rs144421774 | 0 | 2 | 0.4 |  | |  |  |  |
| **DNAH10** | chr12:124415916 | rs200478023 | 0 | 2 | 0.4 |  | |  |  |  |
| **DNAH10** | chr12:124419287 | rs187199487 | 0 | 3 | 0.6 |  | |  |  |  |
| **SRRM2** | chr16:2814476 | rs1438787177 | 0 | 2 | * | 3.78 | | 7.56 | yes | 15.60 |
| **SRRM2** | chr16:2814911 | rs149555495 | 0 | 3 | 0.6 |  | |  |  |  |
| **SRRM2** | chr16:2816519 | rs138447860 | 0 | 6 | 1.19 |  | |  |  |  |
| **SRRM2** | chr16:2816627 | rs117133016 | 1 | 8 | 1.59 |  | |  |  |  |
| **SRRM2** | chr16:2817667 | rs149458303 | 0 | 2 | 0.4 |  | |  |  |  |
| **SIRT1** | chr10:69644532 | chr10:69644532 | 0 | 10 | 1.98 | 2.77 | | 5.54 | yes | 11.40 |
| **SIRT1** | chr10:69672479 | rs116040871 | 0 | 4 | 0.79 |  | |  |  |  |
| **CANT1** | chr17:76989666 | rs34082669 | 0 | 4 | 0.79 | 2.39 | | 4.78 | yes | 9.87 |
| **CANT1** | chr17:76989715 | rs139486406 | 0 | 3 | 0.6 |  | |  |  |  |
| **CANT1** | chr17:76991115 | rs772754957 | 0 | 3 | 0.6 |  | |  |  |  |
| **CANT1** | chr17:76993649 | rs144060377 | 0 | 2 | 0.4 |  | |  |  |  |
| **DPYSL5** | chr2:27121449 | rs139435744 | 0 | 2 | 0.4 | 2.19 | | 4.38 | yes | 9 |
| **DPYSL5** | chr2:27157542 | rs151073506 | 1 | 9 | 1.79 |  | |  |  |  |
| **ABCC10** | chr6:43400205 | rs144459262 | 1 | 8 | 1.59 | 2.19 | | 4.38 | yes | 9 |
| **ABCC10** | chr6:43400850 | rs150124239 | 0 | 3 | * |  | |  |  |  |
| **ABCC10** | chr6:43406366 | rs142854084 | 0 | 3 | 0.6 |  | |  |  |  |
| **DIRC1** | chr2:189599515 | rs148753509 | 0 | 11 | 2.18 | 2.18 | | 4.36 | no |  |
| **ERCC4** | chr16:14029352 | rs41552412 | 0 | 4 | 0.79 | 2.18 | | 4.36 | no |  |
| **ERCC4** | chr16:14029516 | rs1800068 | 0 | 2 | 0.4 |  | |  |  |  |
| **ERCC4** | chr16:14041570 | rs1800069 | 1 | 5 | 0.99 |  | |  |  |  |
| **MCF2L2** | chr3:182923984 | rs61750384 | 0 | 2 | 0.4 | 2 | | 4 | yes | 8.26 |
| **MCF2L2** | chr3:182946129 | rs61753470 | 0 | 3 | 0.6 |  | |  |  |  |
| **MCF2L2** | chr3:183006997 | rs201525425 | 0 | 2 | 0.4 |  | |  |  |  |
| **MCF2L2** | chr3:183017837 | rs144274760 | 0 | 3 | 0.6 |  | |  |  |  |
| **SLC3A1** | chr2:44507990 | rs140317484 | 0 | 2 | 0.4 | 2 | | 4 | no |  |
| **SLC3A1** | chr2:44508526 | rs745458015 | 0 | 3 | 0.6 |  | |  |  |  |
| **SLC3A1** | chr2:44513202 | rs141587158 | 0 | 3 | 0.6 |  | |  |  |  |
| **SLC3A1** | chr2:44539792 | rs121912691 | 0 | 2 | 0.4 |  | |  |  |  |
| **CGNL1** | chr15:57731180 | rs766053332 | 0 | 2 | 0.4 | 2 | | 4 | no |  |
| **CGNL1** | chr15:57820876 | rs144081162 | 0 | 3 | 0.6 |  | |  |  |  |
| **CGNL1** | chr15:57835958 | rs151031750 | 0 | 2 | 0.4 |  | |  |  |  |
| **CGNL1** | chr15:57836791 | rs772155859 | 0 | 2 | 0.4 |  | |  |  |  |
| **APLF** | chr2:68729939 | rs139869963 | 1 | 8 | 1.59 | 1.99 | | 3.98 | no |  |
| **APLF** | chr2:68794471 | rs146949731 | 0 | 2 | 0.4 |  | |  |  |  |
| **MS4A18** | chr11:60496853 | rs140524942 | 0 | 8 | 1.59 | 1.99 | | 3.98 | no |  |
| **MS4A18** | chr11:60497152 | rs76194929 | 0 | 2 | 0.4 |  | |  |  |  |
| **NOD2** | chr16:50745114 | rs104895431 | 0 | 3 | * | 1.99 | | 3.98 | no |  |
| **NOD2** | chr16:50746086 | rs61747625 | 1 | 5 | 0.99 |  | |  |  |  |
| **NOD2** | chr16:50746199 | rs104895444 | 0 | 3 | 0.6 |  | |  |  |  |
| **NOD2** | chr16:50750505 | rs61755272 | 0 | 2 | 0.4 |  | |  |  |  |
| **ELF2** | chr4:139981622 | rs147539143 | 0 | 2 | 0.4 | 1.99 | | 3.98 | yes | 8.22 |
| **ELF2** | chr4:139993175 | rs17322140 | 0 | 8 | 1.59 |  | |  |  |  |
| **DPP9** | chr19:4685696 | rs762729074 | 0 | 2 | 0.4 | 1.98 | | 3.96 | yes | 8.18 |
| **DPP9** | chr19:4689635 | rs200229921 | 0 | 4 | 0.79 |  | |  |  |  |
| **DPP9** | chr19:4704192 | rs200878232 | 0 | 4 | 0.79 |  | |  |  |  |
| **RBM28** | chr7:127953238 | rs73230638 | 1 | 6 | 1.19 | 1.98 | | 3.96 | yes | 8.18 |
| **RBM28** | chr7:127963597 | rs138703329 | 0 | 4 | 0.79 |  | |  |  |  |
| **ZFP91** | chr11:58346953 | rs143372783 | 0 | 6 | 1.19 | 1.98 | | 3.96 | yes |  |
| **ZFP91** | chr11:58347029 | rs200108858 | 0 | 4 | 0.79 |  | |  |  |  |
| **ZSCAN10** | chr16:3139190 | rs185364182 | 0 | 10 | 1.98 | 1.98 | | 3.96 | no |  |
| **ZSCAN10** | chr16:3139999 | rs201495311 | 0 | 10 | * |  | |  |  |  |
| **TMEM175** | chr4:951982 | rs75307864 | 0 | 2 | 0.4 | 1.79 | | 3.58 | yes |  |
| **TMEM175** | chr4:952009 | rs140597786 | 0 | 7 | 1.39 |  | |  |  |  |
| **BABAM2** | chr2:28117455 | rs144572761 | 0 | 9 | 1.79 | 1.79 | | 3.58 | no |  |
| **IPO4** | chr14:24656890 | rs61752845 | 1 | 9 | 1.79 | 1.79 | | 3.58 | no |  |
| **MOCS2** | chr5:52397199 | rs2233218 | 1 | 9 | 1.79 | 1.79 | | 3.58 | yes |  |
| **BICDL2** | chr16:3080997 | rs202141775 | 0 | 4 | 0.79 | 1.78 | | 3.56 | no |  |
| **BICDL2** | chr16:3081048 | rs201144856 | 0 | 5 | 0.99 |  | |  |  |  |
| **CARNS1** | chr11:67186995 | rs41302427 | 0 | 4 | 0.79 | 1.78 | | 3.56 | yes |  |
| **CARNS1** | chr11:67188496 | rs545975688 | 0 | 5 | 0.99 |  | |  |  |  |
| **IGDCC4** | chr15:65682635 | rs185266355 | 0 | 2 | 0.4 | 1.6 | | 3.2 | no |  |
| **IGDCC4** | chr15:65694803 | rs138636327 | 0 | 3 | 0.6 |  | |  |  |  |
| **IGDCC4** | chr15:65703646 | rs142198652 | 0 | 3 | 0.6 |  | |  |  |  |
| **SEMA6B** | chr19:4543938 | rs189240925 | 0 | 2 | 0.4 | 1.59 | | 3.18 | yes |  |
| **SEMA6B** | chr19:4544227 | rs540853448 | 0 | 2 | 0.4 |  | |  |  |  |
| **SEMA6B** | chr19:4558441 | rs773667966 | 0 | 4 | 0.79 |  | |  |  |  |
| **ADGRL2** | chr1:82456107 | rs72719419 | 0 | 4 | 0.79 | 1.59 | | 3.18 | no |  |
| **ADGRL2** | chr1:82456165 | rs143448377 | 0 | 2 | 0.4 |  | |  |  |  |
| **ADGRL2** | chr1:82456341 | rs200383050 | 0 | 2 | 0.4 |  | |  |  |  |
| **ART5** | chr11:3660256 | rs148523079 | 0 | 3 | 0.6 | 1.59 | | 3.18 | no |  |
| **ART5** | chr11:3661129 | rs149052085 | 0 | 5 | 0.99 |  | |  |  |  |
| **KIF7** | chr15:90171738 | rs150248985 | 0 | 5 | 0.99 | 1.59 | | 3.18 | no |  |
| **KIF7** | chr15:90174856 | rs138410949 | 0 | 3 | 0.6 |  | |  |  |  |
| **GNAT2** | chr1:110151344 | rs41280330 | 0 | 8 | 1.59 | 1.59 | | 3.18 | no |  |
| **CALML5** | chr10:5541277 | rs144222183 | 0 | 5 | 0.99 | 1.59 | | 3.18 | no |  |
| **CALML5** | chr10:5541326 | rs61164868 | 0 | 3 | 0.6 |  | |  |  |  |
| **PCSK4** | chr19:1482080 | rs78359732 | 1 | 6 | 1.19 | 1.59 | | 3.18 | no |  |
| **PCSK4** | chr19:1487258 | rs139666108 | 0 | 2 | 0.4 |  | |  |  |  |
| **NID1** | chr1:236176777 | rs148665567 | 0 | 2 | 0.4 | 1.59 | | 3.18 | no |  |
| **NID1** | chr1:236187372 | rs138933538 | 0 | 4 | 0.79 |  | |  |  |  |
| **NID1** | chr1:236228234 | rs147109154 | 0 | 2 | 0.4 |  | |  |  |  |
| **IFI44L** | chr1:79101171 | rs115901054 | 0 | 7 | 1.39 | 1.39 | | 2.78 | no |  |
| **RBL2** | chr16:53495681 | rs61747628 | 0 | 7 | 1.39 | 1.39 | | 2.78 | yes |  |
| **DCBLD2** | chr3:98530317 | rs143662022 | 0 | 4 | 0.79 | 1.39 | | 2.78 | no |  |
| **DCBLD2** | chr3:98536663 | rs149988957 | 0 | 3 | 0.6 |  | |  |  |  |
| **DENND3** | chr8:142170813 | rs142305056 | 0 | 2 | 0.4 | 1.39 | | 2.78 | no |  |
| **DENND3** | chr8:142199128 | rs144442267 | 0 | 5 | 0.99 |  | |  |  |  |
| **HCRTR2** | chr6:55039416 | rs41271312 | 0 | 7 | 1.39 | 1.39 | | 2.78 | no |  |
| **TSR1** | chr17:2235469 | rs62066968 | 0 | 7 | 1.39 | 1.39 | | 2.78 | no |  |
| **ZNF41** | chrX:47307172 | rs144970008 | 0 | 5 | 0.99 | 1.39 | | 2.78 | no |  |
| **ZNF41** | chrX:47307950 | rs766957386 | 0 | 2 | 0.4 |  | |  |  |  |
| **KRT83** | chr12:52709122 | rs148757217 | 0 | 5 | 0.99 | 1.39 | | 2.78 | no |  |
| **KRT83** | chr12:52711549 | rs2857667 | 0 | 2 | 0.4 |  | |  |  |  |
| **AAR2** | chr20:34827932 | rs111591742 | 0 | 3 | 0.6 | 1.2 | | 2.4 | no |  |
| **AAR2** | chr20:34843605 | rs141340897 | 0 | 3 | 0.6 |  | |  |  |  |
| **PHGDH** | chr1:120277956 | rs139063843 | 0 | 2 | 0.4 | 1.2 | | 2.4 | yes |  |
| **PHGDH** | chr1:120279854 | rs149175408 | 0 | 2 | 0.4 |  | |  |  |  |
| **PHGDH** | chr1:120286532 | rs587731325 | 0 | 2 | 0.4 |  | |  |  |  |
| **COQ3** | chr6:99819306 | rs146934336 | 0 | 4 | 0.79 | 1.19 | | 2.38 | yes |  |
| **COQ3** | chr6:99823941 | rs138873880 | 0 | 2 | 0.4 |  | |  |  |  |
| **CRAT** | chr9:131864183 | rs147944652 | 0 | 4 | 0.79 | 1.19 | | 2.38 | no |  |
| **CRAT** | chr9:131864278 | rs372395394 | 0 | 2 | 0.4 |  | |  |  |  |
| **AIFM2** | chr10:71880301 | rs139479025 | 0 | 2 | 0.4 | 1.19 | | 2.38 | no |  |
| **AIFM2** | chr10:71883836 | rs41277978 | 0 | 4 | 0.79 |  | |  |  |  |
| **CNGB3** | chr8:87588042 | rs142846289 | 0 | 4 | 0.79 | 1.19 | | 2.38 | no |  |
| **CNGB3** | chr8:87645092 | rs147876778 | 1 | 2 | 0.4 |  | |  |  |  |
| **CNTD2** | chr19:40730397 | rs972990647 | 0 | 2 | 0.4 | 1.19 | | 2.38 | yes |  |
| **CNTD2** | chr19:40732460 | rs200091528 | 0 | 4 | 0.79 |  | |  |  |  |
| **HGSNAT** | chr8:43054647 | rs112029032 | 0 | 4 | 0.79 | 1.19 | | 2.38 | yes |  |
| **HGSNAT** | chr8:43054684 | rs192857413 | 0 | 2 | 0.4 |  | |  |  |  |
| **IDH2** | chr15:90627553 | rs118053940 | 0 | 4 | 0.79 | 1.19 | | 2.38 | yes |  |
| **IDH2** | chr15:90630704 | rs118101777 | 0 | 2 | 0.4 |  | |  |  |  |
| **RPL3L** | chr16:1997201 | rs370738863 | 0 | 2 | 0.4 | 1.19 | | 2.38 | no |  |
| **RPL3L** | chr16:2004119 | rs146294352 | 0 | 4 | 0.79 |  | |  |  |  |
| **PTPRD** | chr9:8486232 | rs142397137 | 0 | 2 | 0.4 | 1.19 | | 2.38 | yes |  |
| **PTPRD** | chr9:8499805 | rs150444130 | 1 | 4 | 0.79 |  | |  |  |  |
| **TRPM5** | chr11:2437174 | rs111678152 | 0 | 2 | 0.4 | 1.19 | | 2.38 | no |  |
| **TRPM5** | chr11:2437185 | rs34599318 | 0 | 4 | 0.79 |  | |  |  |  |
| **KRTAP12-2** | chr21:46086470 | rs146322780 | 0 | 6 | 1.19 | 1.19 | | 2.38 | no |  |
| **MAGEC2** | chrX:141290950 | rs35094955 | 0 | 6 | 1.19 | 1.19 | | 2.38 | no |  |
| **SLC25A30** | chr13:45970147 | rs138358301 | 0 | 6 | 1.19 | 1.19 | | 2.38 | no |  |
| **CEPT1** | chr1:111690455 | rs41299567 | 0 | 4 | 0.79 | 1.19 | | 2.38 | no |  |
| **CEPT1** | chr1:111690562 | rs752416064 | 0 | 2 | 0.4 |  | |  |  |  |
| **CAMKK2** | chr12:121678569 | rs144382983 | 0 | 2 | 0.4 | 1 | | 2 | yes |  |
| **CAMKK2** | chr12:121678624 | rs1051450715 | 0 | 3 | 0.6 |  | |  |  |  |
| **ACOT12** | chr5:80628348 | rs143972820 | 0 | 2 | 0.4 | 1 | | 2 | no |  |
| **ACOT12** | chr5:80640844 | rs147070236 | 0 | 3 | 0.6 |  | |  |  |  |
| **ECM2** | chr9:95272297 | rs41278707 | 0 | 3 | 0.6 | 1 | | 2 | no |  |
| **ECM2** | chr9:95279986 | rs147710026 | 0 | 2 | 0.4 |  | |  |  |  |
| **MFSD12** | chr19:3544812 | rs200371465 | 1 | 2 | 0.4 | 1 | | 2 | yes |  |
| **MFSD12** | chr19:3546264 | rs34878396 | 0 | 3 | 0.6 |  | |  |  |  |
| **MINK1** | chr17:4794407 | rs200592676 | 0 | 3 | 0.6 | 1 | | 2 | yes |  |
| **MINK1** | chr17:4794973 | rs199633382 | 0 | 2 | 0.4 |  | |  |  |  |
| **NF1** | chr17:29496980 | chr17:29496980 | 0 | 2 | 0.4 | 1 | | 2 | yes |  |
| **NF1** | chr17:29496992 | chr17:29496992 | 0 | 3 | 0.6 |  | |  |  |  |
| **TLR3** | chr4:187003729 | rs35311343 | 0 | 3 | 0.6 | 1 | | 2 | no |  |
| **TLR3** | chr4:187005935 | rs146846379 | 0 | 2 | 0.4 |  | |  |  |  |
| **UBASH3B** | chr11:122526821 | rs150696870 | 0 | 3 | 0.6 | 1 | | 2 | yes |  |
| **UBASH3B** | chr11:122680567 | rs139627911 | 0 | 2 | 0.4 |  | |  |  |  |
| **ANOS1** | chrX:8503715 | rs137900287 | 0 | 5 | 0.99 | 0.99 | | 1.98 | no |  |
| **GPR132** | chr14:105517675 | rs61737948 | 0 | 5 | 0.99 | 0.99 | | 1.98 | no |  |
| **ENPP1** | chr6:132172374 | rs142001296 | 0 | 5 | 0.99 | 0.99 | | 1.98 | no |  |
| **RHBDF1** | chr16:109431 | rs764126066 | 0 | 1 | 0.2 | 0.99 | | 1.98 | no |  |
| **RHBDF1** | chr16:111141 | rs976221912 | 0 | 4 | 0.79 |  | |  |  |  |
| **KCNH8** | chr3:19479731 | rs138531032 | 0 | 5 | 0.99 | 0.99 | | 1.98 | no |  |
| **CSF3R** | chr1:36933715 | rs3917996 | 0 | 2 | 0.4 | 0.8 | | 1.6 | no |  |
| **CSF3R** | chr1:36937710 | rs369185176 | 0 | 2 | 0.4 |  | |  |  |  |
| **FRMD3** | chr9:85863069 | rs190298518 | 0 | 2 | 0.4 | 0.8 | | 1.6 | no |  |
| **FRMD3** | chr9:85863101 | rs200181003 | 0 | 2 | 0.4 |  | |  |  |  |
| **ENPP3** | chr6:131962602 | rs41285342 | 0 | 2 | 0.4 | 0.8 | | 1.6 | no |  |
| **ENPP3** | chr6:131962632 | rs41286144 | 0 | 2 | 0.4 |  | |  |  |  |
| **KRT12** | chr17:39020022 | rs139647784 | 0 | 2 | 0.4 | 0.8 | | 1.6 | no |  |
| **KRT12** | chr17:39022440 | rs147584242 | 0 | 2 | 0.4 |  | |  |  |  |
| **PHC1** | chr12:9085452 | rs12810327 | 0 | 2 | 0.4 | 0.8 | | 1.6 | no |  |
| **PHC1** | chr12:9090612 | rs183583604 | 0 | 2 | 0.4 |  | |  |  |  |
| **SLC34A3** | chr9:140130522 | rs138872455 | 0 | 2 | 0.4 | 0.8 | | 1.6 | no |  |
| **SLC34A3** | chr9:140130795 | rs200090657 | 0 | 2 | 0.4 |  | |  |  |  |
| **TRIM32** | chr9:119460579 | rs117599771 | 0 | 2 | 0.4 | 0.8 | | 1.6 | no |  |
| **TRIM32** | chr9:119461243 | rs3747835 | 0 | 2 | 0.4 |  | |  |  |  |
| **ZFHX4** | chr8:77764484 | rs139920573 | 1 | 4 | 0.79 | 0.79 | | 1.58 | no |  |
| **RASSF1** | chr3:50368871 | rs150999288 | 0 | 4 | 0.79 | 0.79 | | 1.58 | no |  |
| **RASSF1** | chr3:50369058 | rs148222115 | 0 | 2 | * |  | |  |  |  |
| **COL4A4** | chr2:227924228 | rs36121515 | 0 | 4 | 0.79 | 0.79 | | 1.58 | no |  |
| **SLC9A8** | chr20:48429485 | rs61734269 | 0 | 3 | 0.6 | 0.6 | | 1.2 | no |  |
| **PNPLA2** | chr11:824567 | rs202081894 | 0 | 3 | 0.6 | 0.6 | | 1.2 | no |  |
| **STARD8** | chrX:67937939 | rs151192738 | 0 | 1 | 0.2 | 0.6 | | 1.2 | no |  |
| **STARD8** | chrX:67941903 | rs138190419 | 0 | 1 | 0.2 |  | |  |  |  |
| **STARD8** | chrX:67942274 | chrX:67942274 | 0 | 1 | 0.2 |  | |  |  |  |
| **NUP155** | chr5:37331792 | rs137866662 | 0 | 3 | 0.6 | 0.6 | | 1.2 | no |  |
| **WWC2** | chr4:184182125 | rs141501417 | 0 | 3 | 0.6 | 0.6 | | 1.2 | no |  |
| **USP17L13** | chr4:9227402 | chr4:9227402 | 0 | 3 | 0.6 | 0.6 | | 1.2 | no |  |
| **USP17L13** | chr4:9227418 | chr4:9227418 | 0 | 3 | * |  | |  |  |  |
| **HEPH** | chrX:65412133 | chrX:65412133 | 0 | 1 | 0.2 | 0.6 | | 1.2 | no |  |
| **HEPH** | chrX:65417554 | rs151003259 | 0 | 2 | 0.4 |  | |  |  |  |
| **SNTG2** | chr2:1271281 | rs201124856 | 0 | 2 | 0.4 | 0.4 | | 0.8 | no |  |
| **SPNS2** | chr17:4439574 | rs768133723 | 0 | 2 | 0.4 | 0.4 | | 0.8 | yes |  |
| **ST18** | chr8:53077749 | rs34326988 | 0 | 2 | 0.4 | 0.4 | | 0.8 | yes |  |
| **SV2C** | chr5:75596612 | rs188730765 | 0 | 2 | 0.4 | 0.4 | | 0.8 | yes |  |
| **WDR35** | chr2:20166496 | rs143343508 | 0 | 2 | 0.4 | 0.4 | | 0.8 | no |  |
| **ADAMTS19** | chr5:128861979 | rs758981800 | 0 | 2 | 0.4 | 0.4 | | 0.8 | no |  |
| **APC2** | chr19:1468408 | rs200444154 | 0 | 2 | 0.4 | 0.4 | | 0.8 | yes |  |
| **GLIS2** | chr16:4387209 | rs75495782 | 0 | 2 | 0.4 | 0.4 | | 0.8 | no |  |
| **KIAA1109** | chr4:123167847 | rs75923324 | 0 | 2 | 0.4 | 0.4 | | 0.8 | yes |  |

*inf: variants present in more than one individual

**Supplementary Table 5: Table of individuals in the SW cohort carrying two or more variants within the top 10 genes.**

The table shows the number of rare variants per sample across these genes

| **Patient** | **NPIPB13** | **SIRT1** | **SRRM2** | **CANT1** | **DPYSL5** | **ABCC10** | **MCF2L2** | **ELF2** | **DPP9** | **RBM28** | **Variant Description** |
| --- | --- | --- | --- | --- | --- | --- | --- | --- | --- | --- | --- |
| **Patient 1** | **x** | **x** |  |  |  |  |  |  |  |  | NPIPB13 (rs796810494); SIRT1 (chr10:69644532) |
| **Patient 2** | **x** |  |  |  |  |  |  | **x** |  |  | NPIPB13 (rs796810494); ELF2 (rs17322140) |
| **Patient 3** | **x** |  | **x** |  |  |  |  |  |  |  | NPIPB13 (rs796810494); SRRM2 (rs1438787177, rs117133016) |
| **Patient 4** | **x** | **x** |  |  |  |  |  |  |  | **x** | NPIPB13 (rs796810494); SIRT1 (chr10:69644532); RBM28 (rs73230638) |
| **Patient 5** | **x** |  |  |  |  |  |  | **x** |  |  | NPIPB13 (rs796810494); ELF2 (rs200074545) |
| **Patient 6** | **x** |  |  |  |  |  |  |  |  | **x** | NPIPB13 (rs796810494); RBM28 (rs73230638) |
| **Patient 7** | **x** |  |  | **x** |  |  |  |  |  |  | NPIPB13 (rs796810494); CANT1 (rs34082669) |
| **Patient 8** | **x** |  |  | **x** |  |  |  |  |  |  | NPIPB13 (rs796810494); CANT1 (rs144060377) |
| **Patient 9** | **x** |  |  | **x** |  |  | **x** |  |  |  | NPIPB13 (rs796810494); CANT1 (rs144060377), MCF2L2 (rs144274760) |
| **Patient 10** | **x** |  |  |  |  |  |  |  |  | **x** | NPIPB13 (rs796810494); RBM28 (rs73230638) |
| **Patient 11** | **x** |  | **x** |  |  |  |  |  |  |  | NPIPB13 (rs796810494); SRRM2 (rs1438787177) |
| **Patient 12** | **x** | **x** |  |  |  |  |  |  |  |  | NPIPB13 (rs796810494); SIRT1 (rs116040871) |
| **Patient 13** | **x** |  | **x** |  | **x** |  |  |  | **x** |  | NPIPB13 (rs1240720619); SRRM2 (rs117133016); DPYSL5 (rs151073506); DPP9 (rs200878232) |
| **Patient 14** |  | **x** | **x** |  |  |  |  |  | **x** |  | SIRT1 (chr10:69644532); SRRM2 (rs138447860); DPP9 (rs200878232) |
| **Patient 15** |  | **x** | **x** |  | **x** |  |  |  |  |  | SIRT1 (rs116040871); SRRM2 (rs117133016); DPYSL5 (rs139435744) |
| **Patient 16** |  | **x** | **x** |  |  |  |  |  |  |  | SIRT1 (chr10:69644532); SRRM2 (rs117133016) |
| **Patient 17** |  |  | **x** |  | **x** |  |  |  |  |  | SRRM2 (rs117133016); DPYSL5 (rs151073506) |
| **Patient 18** |  | **x** | **x** |  |  |  |  |  |  |  | SRRM2 (rs138447860); SIRT1 (rs548590752) |
| **Patient 19** |  |  | **x** |  | **x** | **x** |  |  |  |  | SRRM2 (rs138447860); DPYSL5 (rs139435744); ABCC10 (rs144459262) |
| **Patient 20** |  |  | **x** |  |  | **x** |  | **x** |  |  | SRRM2 (rs138447860); ABCC10 (rs144459262); ELF2 (rs17322140) |
| **Patient 21** |  |  | **x** |  |  |  |  | **x** |  | **x** | SRRM2 (rs149555495); ELF2 (rs147539143); RBM28 (rs138703329) |
| **Patient 22** |  |  |  |  | **x** |  | **x** |  |  |  | DPYSL5 (rs151073506); MCF2L2 (rs61753470) |
| **Patient 23** |  |  |  |  |  | **x** |  |  | **x** |  | ABCC10 (rs142854084); DPP9 (rs200878232) |
| **Patient 24** |  | **x** |  |  |  |  | **x** |  |  |  | SIRT1 (chr10:69644532); MCF2L2 (rs61750384) |
